# Exploring the neuromagnetic signatures of cognitive decline from mild cognitive impairment to Alzheimer’s disease dementia

**DOI:** 10.1101/2024.07.06.24310016

**Authors:** Sinead Gaubert, Pilar Garces, Jörg Hipp, Ricardo Bruña, Maria Eugenia Lopéz, Fernando Maestu, Delshad Vaghari, Richard Henson, Claire Paquet, Denis Engemann

## Abstract

**Introduction:** Alzheimer’s disease (AD) is the most common cause of dementia. Non-invasive, affordable, and largely available biomarkers that are able to identify patients at a prodromal stage of AD are becoming essential, especially in the context of new disease-modifying therapies. Mild cognitive impairment (MCI) is a critical stage preceding dementia, but not all MCI patients will progress to AD. This study explores the potential of non-invasive magnetoencephalography (MEG) to predict future cognitive decline from MCI to AD dementia.

**Methods:** We analyzed resting state MEG data from the BioFIND dataset including 117 MCI patients, of whom 64 progressed to AD dementia (AD progression) while 53 remained stable (stable MCI) using multivariate spectral analyses. The patients were followed-up between 2009 and 2018. Receiver operating characteristic curves obtained via logistic regression models were used to quantify separation of patients progressing to AD dementia from stable MCI.

**Results:** MEG beta power, particularly over parieto-occipital magnetometers, was significantly reduced in the AD progression group compared to stable MCI, indicative of future cognitive decline. Logistic regression models showed that MEG beta power outperformed conventional metrics like the Mini Mental Status Examination (MMSE) score and structural brain measures in predicting progression to AD dementia (AUC 0.81 vs 0.71 and AUC 0.81 vs 0.75, respectively). The combination of age, education, MMSE, MEG beta power and Hippocampal volume/Total grey matter ratio achieved a 0.83 AUC, 78% sensitivity and 76% specificity. Spectral covariance matrices analyzed with Riemannian methods exhibited significant differences between groups across a wider range of frequencies than spectral power.

**Discussion:** These findings highlight the potential of spectral power and covariance as robust non-invasive electrophysiological biomarkers to predict MCI progression that complement other diagnostic measures, including cognitive scores, structural magnetic resonance imaging (MRI) and biological biomarkers.

## Introduction

Alzheimer’s disease (AD) is the most common cause of dementia, accounting for an estimated 60% to 80% of cases (Alzheimer’s Association, 2023). Dementia is preceded by the mild cognitive impairment (MCI) condition, which is characterized by objective cognitive impairment in one or more domains with preserved functional independence (Petersen et al., 2014). Not all MCI patients will progress to AD dementia, as some can stay cognitively stable or even revert to normal cognition (Hampel & Lista, 2016). Recently, new AD treatments have been developed with promising results (Sims et al., 2023; Van Dyck et al., 2023). Biomarkers that are able to identify patients at a prodromal stage of AD are becoming essential, as treatments have shown to be more effective if given at an early stage. Biomarkers of AD progression are also needed to monitor the response to new treatments. However, many AD biomarkers either require an invasive procedure (cerebrospinal fluid analysis), or are associated with high costs and limited availability, such as amyloid-PET and Tau-PET scans, so they cannot be applied to a large population sample worldwide, especially with repeated measures. Specifically, AD biomarkers (beta-amyloid protein, tau and phosphorylated tau that can be measured in cerebrospinal fluid, plasma or positron emission tomography) have a relatively low sensitivity to synaptic dysfunction and may be insufficient to monitor modifications of brain function under treatment (Jack et al., 2018; Janelidze et al., 2023; Li et al., 2022; Thijssen et al., 2021). Moreover, existing biomarkers do not account for the disjunction between the degree of brain pathology and its clinical manifestations, which refers to the concept of cognitive reserve (Stern, 2009). It has been shown that individuals with similar brain pathology can demonstrate differences in cognitive performance, probably underpinned by variations in functional network efficiency (Balart-Sánchez et al., 2021; Ewers et al., 2021; Lee et al., 2022). This indicates an unmet need for brain activity-based biomarkers.

Non-invasive electrophysiology, such as electroencephalography (EEG) and magnetoencephalography (MEG), are promising techniques that could be complementary to other biomarkers currently in development for AD, including biological and imaging biomarkers (Dauwels et al., 2010; Engemann et al., 2020; Gaubert et al., 2019; Stam, 2010). Electrophysiology allows for the examination of neuronal activity across spatial and temporal scales, providing a window onto neuronal activity underlying cognitive functioning with high sensitivity to synaptic function (Babiloni et al., 2006; Schnitzler & Gross, 2005). Information can be decoded from M/EEG by analyzing the spatial and spectral organization of brain activity using advanced statistical methods including machine learning (King & Dehaene, 2014; Stam et al., 2003; Stokes et al., 2015). EEG and MEG have differential sensitivity to different configurations of neural activity (e.g, in terms of the orientation and depth of the dendritic currents that cause the electromagnetic field changes, and the effects of volume conduction and skull/scalp conductivities). While EEG is more suitable for clinical deployment due to better standardisation, scalability and costs, MEG offers better spatial resolution and often higher signal-to-noise ratio, making it a useful tool for research-grade discovery contexts (Babiloni et al., 2020; Lehtelä et al., 1997). Moreover, recent developments in MEG sensors, like optically-pumped magnetometers (Brookes et al., 2022), promise better scalability and reduced cost, i.e., more practical applications in the clinic. Despite some intrinsic differences between MEG and EEG, under favourable circumstances (e.g. spectral pattern of limited spatial complexity), MEG signatures could also be validated and adapted for biomarker studies using EEG (Frey et al., 2014; Weisz et al., 2014; Wöstmann et al., 2019). Although MEG is still undervalued in AD, it has the potential to significantly contribute to our understanding of how neurodegenerative diseases impact brain function, and could help predict future cognitive decline (Benwell et al., 2020; Osipova et al., 2005).

A solid body of evidence exists that characterizes temporal/spectral differences in resting-state brain activity between healthy controls and patients with MCI or AD dementia, as assessed with EEG. As reviewed by Cassani et al. (2018), compared to healthy controls, patients with AD or MCI usually show: 1) slowing of oscillatory brain activity, which is thought to result from loss of cholinergic innervation; 2) reduced signal complexity, which could be linked to neurodegeneration and fewer cortical connections; and 3) reduced synchrony, which likely reflects impaired communication of neural networks. While previous studies have used EEG/MEG in the context of MCI and AD dementia versus controls (e.g., Babiloni et al., 2009, 2010, 2016; Blinowska et al., 2017; Dauwels et al., 2010; Garcés et al., 2013; Jeong, 2004; Wen et al., 2015; Vaghari et al., 2022b), fewer studies have used EEG to predict progression from MCI to AD dementia (e.g. Engedal et al., 2020; Moretti, 2015; Musaeus et al., 2020; Poil et al., 2013; Rossini et al., 2006) and even fewer have used MEG for this purpose (Fernández et al., 2006; López et al., 2014; Maestú & Fernández, 2020). Moreover, previous work has mostly focussed on specific features (e.g. power in specific frequency bands), which also differ between studies, rendering comparisons difficult. Furthermore, pathology and medical treatments may alter neural dynamics at frequencies that are not well represented by standard frequency bands (Jobert & Wilson, 2015). This motivates approaches that analyze the frequency spectrum continuously (Hawellek et al., 2022; Hipp et al., 2012), particularly since different pathological conditions can modulate electrophysiological signals at different spatial scales (Bomatter et al., 2023). For example, oscillatory activity in the alpha band (∼10Hz) tends to be maximal over posterior visual cortices, while that in the beta band (∼20Hz) tends to be maximal over motor cortex (Bourguignon et al., 2019). In addition to changes in power, there can be changes in phase-coupling mechanisms (Aydore et al., 2013; Vinck et al., 2011) and slow fluctuations of amplitude envelopes (Brookes et al., 2012; Hipp et al., 2012), as part of large-scale cortical network dynamics (Siegel et al., 2012).

This leads to the following two scientific questions for the present study: 1) can brain activity recorded with MEG help predict progression from MCI to AD dementia, and 2) what features of the MEG signal are most characteristic of future cognitive decline? To address these questions, we analyzed neuromagnetic recordings from 117 MCI patients, of which 64 later progressed to AD dementia (AD progression) while 53 remained cognitively stable (stable MCI) within 9-year follow-up, using 306-channel wholehead MEG. To avoid bias due to pre-specified frequency bands, we conducted continuous spectral analysis using Morlet wavelets, covering the frequency spectrum from 1 Hz to 64 Hz in fine-grained intervals. This allowed us to define a common signal representation for various spectral measures used in the literature, i.e., power, covariance, and synchronization measures including phase interactions, and power envelopes. As fine-grained regional changes in cortical activity might be lost when averaging across sensors, we employed multivariate analysis using the mathematical framework of Riemannian manifolds. These tools are well suited for capturing fine-grained spatio-spectral patterns of brain activity that are confounded by volume conduction and field spread – bypassing the need for source localization with a biophysical model.

## Methods

### Participants

We analyzed MEG recordings from the BioFIND dataset (Vaghari et al., 2022a) comprising 158 clinically diagnosed MCI individuals according to Albert et al. (2011) criteria, recruited from two sites: 68 patients from the MRC Cognition & Brain Sciences Unit (CBU) at the University of Cambridge and 90 patients from the Laboratory of Cognitive and Computational Neuroscience at the Centre for Biomedical Technology (CTB), Madrid. The participants were pooled over several different studies, each approved by local Ethics Committees and following the 1991 Declaration of Helsinki.

We used a subset of 117 of the MCI patients in BioFIND who had follow-up data. Of these, 64 subsequently progressed to probable AD dementia based on clinical criteria according to McKhann et al. (2011) (AD progression group), whereas 53 remained stable (stable MCI). For additional details, see the BioFIND dataset publication (Vaghari et al., 2022a).

### MEG data acquisition

MEG recordings were collected continuously at 1 kHz sample rate using an Elekta Neuromag Vectorview 306 MEG system (Helsinki, FI) at both CBU and CTB sites. Resting-state MEG data were recorded while participants were seated comfortably inside a magnetically shielded room and were asked to keep their eyes closed, but not fall asleep.

### Preprocessing

We analyzed the first 2 minutes of resting-state eyes-closed MEG data for each patient. Data were processed in Python using the MNE software version 1.2.0 (Gramfort et al., 2013). The preprocessing steps were the following: MaxFiltering (SSS) was first applied to raw data to remove noise potentially arising from head movements and environmental noise. We applied the MaxFilter process using site-specific calibration and cross-talk correction files, as used in the study by Vaghari et al. (2022a). Temporal Signal Space Separation (tSSS) was employed with an st_duration parameter of 10 seconds to enhance the separation of brain and external signals. The head origin was automatically determined (mf_head_origin = ‘auto’), and the destination was set to (dev_head_t), ensuring consistent spatial alignment despite potential head movements. Data was resampled to a rate of 250 Hz, after which a 0.5 Hz to 100 Hz 4th-order Butterworth bandpass filter and 50 Hz Notch filter were applied. To remove ocular and cardiac artifacts, spatial filtering was employed using the *signal space projection* (SSP) technique (Uusitalo & Ilmoniemi, 1997). The data were then cut in 10-second epochs and the *autoreject* algorithm was used to exclude noisy epochs (Jas et al., 2017).

### Computation of MEG features

We focused on magnetometers as after MaxFilter cleaning, the information of gradiometers and magnetometers is merged and duplicated across both sensor types. Indeed, previous work has shown that after MaxFilter cleaning, spectral results obtained from gradiometers and magnetometers are highly similar (Garcés et al., 2017) and comparison of the power spectra between gradiometers and magnetometers on this dataset led us to the same conclusion. This facilitated data analysis through simpler handling and shorter computation times.

We computed spectral features with Morlet wavelets (Hipp et al., 2012) using the meeglet library (Bomatter et al., 2023). This wavelet approach implements Morlet wavelets spaced on a logarithmic grid such that the spacing between wavelets and their spectral smoothness increase log-linearly with frequency (Bomatter et al., 2023; Hipp et al., 2012). Such wavelets are well suited for capturing the log-dynamic frequency scaling of brain activity (Buzsáki & Mizuseki, 2014) and have been proven useful in multiple EEG-biomarker applications (Frohlich et al., 2019; Hawellek et al., 2022; Hipp et al., 2021). Moreover, Bomatter and colleagues found that such wavelets could outperform classical frequency-band definitions or linearly spaced power spectra. Another advantage of this approach is that across various spectral measures, the same spectral analysis method is used in this work, reducing methods-induced variance. The frequency of interest ranged from 1 Hz to 64 Hz, with a spectral smoothing (bandwidth) of 0.35 oct and a spectral sampling of 0.05 oct. The bandwidth was chosen based on visual inspection of the average power spectrum across participants to control the trade-off between smoothness and spectral resolution.

The following spectral features were computed: power spectral density, covariance estimated from the wavelet-convoluted timeseries, debiased squared weighted phase-lag index (dwPLI) and power envelope correlation (log of rectified wavelet-convoluted timeseries). This set of spectral features can capture distinct aspects of neural activity and are complementary. Previous EEG work has shown that spectral power, dwPLI and power envelope correlation revealed complementary facets of brain function in Huntington’s Disease and pharmacological treatments thereof (Hawellek et al., 2022). Spectral power density quantifies frequency-specific brain activity. The covariance provides an extension of spectral power as it includes the power spectrum but also provides information about the correlation between sensors. This can help unmix hidden activity patterns and assess interdependence of neural signals: covariance-based modelling has recently been explored in machine learning to uncover brain activity without explicit MRI-based source localization (Sabbagh et al., 2020). On the other hand, the dwPLI metric captures changes in phase-synchronization with reduced sensitivity to uncorrelated noise sources and increased statistical power to detect changes in phase-synchronization compared to PLI (Vinck et al., 2011). Power envelope correlation can detect non-instantaneous power correlations - regardless of their sign – which has been used to study synchronised signal amplitude changes between distant brain regions (Hipp et al., 2012), The power envelope was defined as the log of the rectified wavelet-convoluted signal (Bomatter et al., 2023; Hipp et al., 2012).

Together, these metrics provide complementary information on brain activity in different frequency ranges. We were in particular interested in studying metrics that can be defined in sensor space, and therefore more easily transposed to clinical settings (where an MRI may not be available, for example, for the accurate head modelling needed for MEG/EEG source inversion). In this work, we therefore refrained from an interpretation of these metrics in terms of functional connectivity, which for proper interpretation requires anatomical source modelling.

### MRI features

To assess the complementarity of MEG signals to anatomical information, we analyzed T1-weighted structural MRIs from the BioFIND dataset. T1-weighted MRIs were processed using FreeSurfer software (Fischl, 2012) to compute global brain volumetric measures. Based on the previous AD literature (Hu et al., 2023; Rathore et al., 2017), we focused on two specific brain volumetric measures that have been demonstrated as highly relevant for predicting AD progression: log ratio of mean hippocampal volume to total grey matter, and log of the lateral ventricle volume. Reduced hippocampal volume is a well-established marker for AD (Jack et al., 2018), where its normalization relative to total grey matter volume helps to account for individual differences in total brain size. Increased ventricular volume is associated with brain atrophy and is also considered a marker relevant to AD progression (Marizzoni et al., 2019).

### Statistical analysis

We compared socio-demographic characteristics between the two MCI groups using Student’s t-test for continuous variables and χ2 test for categorical variables.

#### Uncorrected inference by frequency

For spectral power analysis, we computed and plotted the average log power spectra over all sensors between 1Hz and 64Hz. We used non-parametric bootstrap resampling to obtain confidence intervals and permutation tests of the mean difference to obtain (uncorrected) p-values for the plotted average difference between groups along the frequency spectrum. Both were implemented using Scipy’s (Virtanen et al., 2020) `bootstrap` and `permutation_test` functions, respectively, with 9999 iterations (default).

#### Clustering permutation-testing across frequencies

To correct for multiple comparisons and take into account the correlation between frequencies, we used clustering-permutation tests along the frequency spectrum (permutation F-test) as implemented in MNE Python (Gramfort et al., 2013).

The permutation F-test was used to compare average spectral power between groups at frequencies ranging from 1 to 64 Hz. For power envelope correlation and dwPLI, we computed the average metrics over all sensors. A permutation F-test was used to compare average metrics between groups at frequencies ranging from 1 Hz to 64 Hz, using an F-test at each frequency followed by clustering and permutation. We used 10000 iterations.

#### Clustering permutation-testing across frequencies and sensors

For each metric (spectral power, dwPLI and power envelope correlation), we computed topographical maps containing the metric of interest for every sensor and each group. To test for regional differences, we computed spatial-spectral permutation tests adapted from MNE-Pythons spatio-temporal permutation testing procedures. To reduce the dependency of the results on the particular choice of cluster-inclusion values, we used Threshold-Free Cluster Enhancement (TFCE) as implemented by the MNE-Python software (Smith & Nichols, 2009). As this procedure was more computationally costly, we used 1024 permutations (default in MNE).

#### Covariance-based Riemannian distance MANOVA

As a complementary method for using spatial information in statistical analysis, we explored multivariate Riemannian distance MANOVA F-test (Anderson, 2001) implemented in the Pyriemann library (Barachant et al., 2023). This procedure conducts non-parametric distance MANOVA to test for group differences (Anderson, 2001). As a distance, we used the Riemannian affine invariant distance which has been used with great success in different machine learning applications for MEG and EEG including brain computer interfaces, brain-age prediction and sex classification (Barachant et al., 2012; Bomatter et al., 2023; Congedo et al., 2017; Sabbagh et al., 2020). Theoretical analysis and empirical benchmarks have shown that Riemannian metrics mitigate distortions due to electromagnetic field spread and provide latent representations well suited to statistically isolate information related to cortical current generators (Sabbagh et al., 2019, 2020). Combined with the distance MANOVA, this procedure can be expected to provide a useful multivariate method for detecting group differences in brain activity. The affine-invariant Riemannian metric expects covariances to be symmetric positive definite (SPD), hence, to be full rank. To obtain valid SPD matrices, we used the method from Sabbagh et al. (2019), which linearly projects covariance matrices to the smallest common subspace using principal component analysis. As Maxfiltering projects noise components from the data and commonly reduces the data rank to 65, we projected the covariances to the rank of 65 and applied a regularization parameter of 1e-15.

#### Logistic regression model of progression risk

Finally, we implemented logistic regression models in R Software Version 2022.12.0+353 to assess the relationship between several predictors (demographical data, MMSE, MEG spectral power and brain MRI volumes) and the probability of progression to AD dementia. We employed model selection criteria, specifically AIC (Akaike Information Criterion), to identify the most appropriate model in terms of trading expected generalization error against model complexity. We computed marginal effects to perform inference in terms of implied changes in probability, rather than the linear predictor of the generalized linear model (Leeper, 2017). Marginal effects were computed alongside with performance metrics (Area Under the Curve, Sensitivity, Specificity).

All analysis code used in this study will be made available on GitHub upon publication of this manuscript.

## Results

We analyzed resting state MEG data from 117 MCI patients, of whom 64 progressed to AD dementia (AD progression) while 53 remained stable (stable MCI).

### Socio-demographic characteristics

There were no significant differences for AD progression group and stable MCI for age or sex. There was a trend for a difference in years of education, though this actually suggested longer education in the AD progression group, suggesting that progression did not simply reflect worse education (**Table 1).** The Mini-Mental Status Examination (MMSE) score was significantly lower in patients showing AD progression compared to stable MCI (p=0.004) – see Discussion.

**Table 1:** *Means (and standard deviations) of socio-demographic characteristics of AD progression group and stable MCI.* M: Male; F: Female; MMSE: Mini Mental State Examination.

| Data Characteristic | AD progression (n=64) | Stable MCI (n=53) | T/$\chi^2$ and p-value |
| --- | --- | --- | --- |
| Sex (M/F) | 32/32 | 20/33 | $\chi^2 = 1.45$, $p=0.23$ |
| Age (years) | 72.96 (7.21) | 72.60 (5.42) | T= -0.29, $p=0.77$ |
| Education (years) | 10.28 (4.53) | 8.56 (4.63) | T = -1.92, $p=0.06$ |
| Baseline MMSE | 25.60 (2.96) | 27.16 (2.38) | T= 2.97, $p=0.004$ |

### Analysis of frequency-dependent brain activity

AD progression was associated with reduced averaged spectral power compared to stable MCI in frequencies ranging from 10 Hz to 50 Hz (**Figure 1 A-C**). Topographical analysis showed reduced spectral power over posterior sensors for AD progression in frequencies from 9.5 Hz to 62 Hz (**Figure 1E**). As a sensitivity analysis, adjustment on MMSE was done by residualizing the metric of interest using a linear regression model. After adjustment on MMSE, significant group differences in spectral power remained, with the AD progression group showing reduced 16 Hz to 36 Hz power over left parieto-occipital sensors (**Supplementary Figure 1**). An analysis based on the full covariance matrix using Riemannian-distance MANOVA F-test confirmed the high-frequency difference and, in addition, uncovered significant group differences in frequencies from 1Hz to 4Hz (**Figure 1D**).

**Figure 1:**
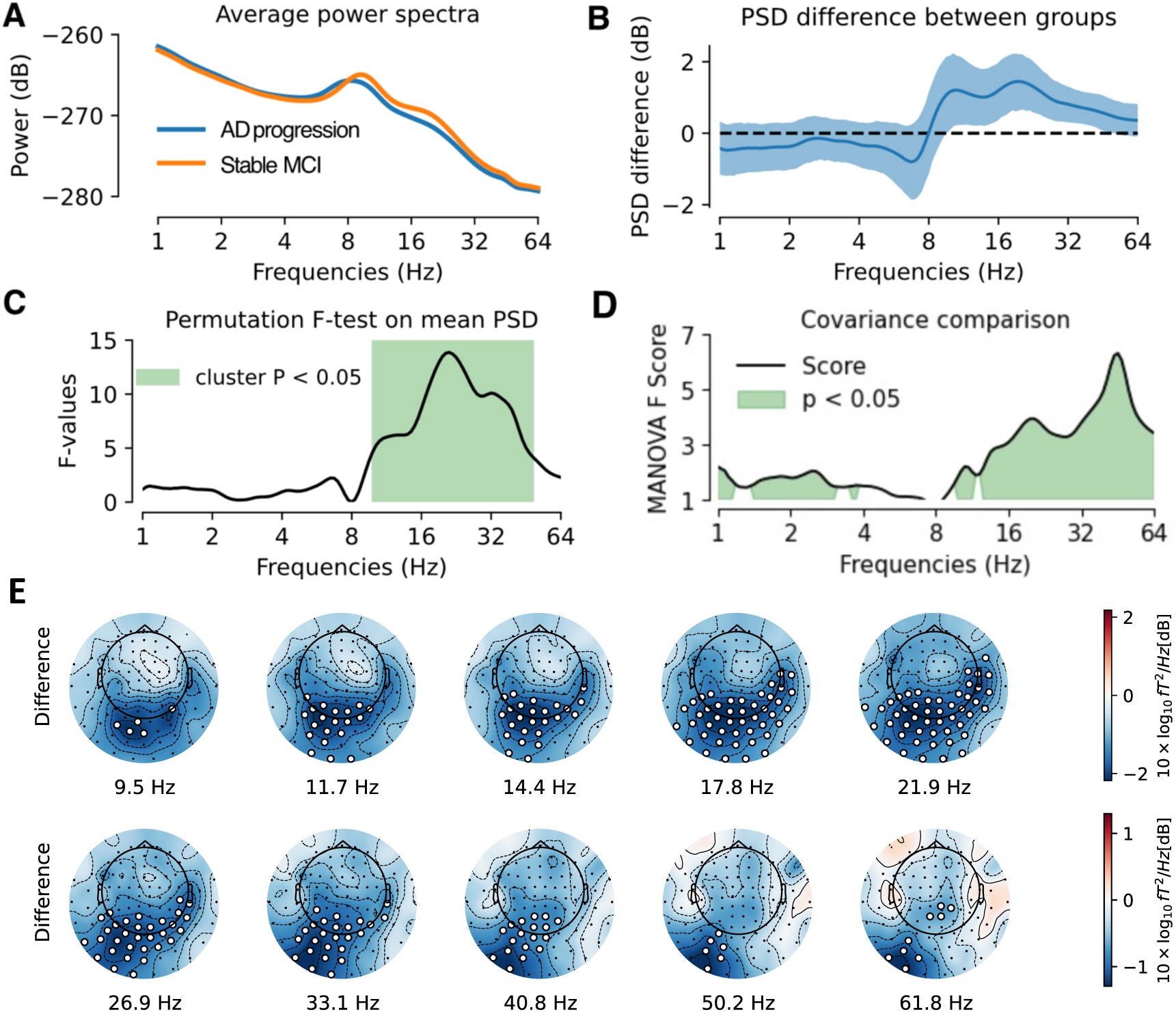
(**A)** Power spectra averaged over all sensors: AD progression was associated with reduced spectral power at baseline in frequencies ranging from 10 Hz to 50 Hz. (**B)** Mean spectral power difference between groups (blue line) and 95% confidence interval computed by bootstrap (blue shaded area). (**C)** Permutation F-test on mean spectral power, showing significant power difference between AD progression and stable MCI at frequencies ranging from 10 Hz to 50 Hz. (**D)** Comparison between groups based on frequency-specific covariances using Riemannian-distance MANOVA, showing significant differences between 1 Hz to 4 Hz and 10 Hz to 64 Hz. (**E)** Topographical maps of spectral power difference between groups, showing reduced spectral power over posterior sensors in AD progression group in frequencies from 9.5 Hz to 62 Hz. The white dots indicate significant differences (p < 0.05).

### Analysis of phase and amplitude interactions

We next explored the presence of informative differences in measures associated with long-range neural interactions beyond the power spectrum. Visual inspection suggested that AD progression was associated with reduced power envelope correlation and reduced dwPLI around 8 Hz. However, results were not statistically significant following permutation F-test with multiple-comparison correction across all frequencies. In the topographical analysis, the AD progression group showed a spatially consistent pattern of decreased dwPLI and power envelope correlation in the alpha frequency band over posterior sensors, but this did not survive correction for multiple comparisons (**Supplementary Figure 2 and Supplementary Figure 3**).

### Multimodal analysis of AD progression risk

We computed logistic regression models to assess the relationship between several predictors (age, sex, education, MMSE, MEG spectral power, log ratio of hippocampal volume to total grey matter and log lateral ventricle volume) and the probability of progression to AD dementia (**Figure 2 and Supplementary Figure 4**). For MEG power, we used the cluster values obtained after adjustment for MMSE from our previous analysis, e.g., spectral power between 16 Hz and 36 Hz in left parieto-occipital sensors. Based on AIC values, the favored model combined age, education, MMSE, cluster MEG beta power and Hippocampus/Total grey matter ratio (**Figure 2**). Lower values of MEG 16Hz to 36Hz power in left parieto-occipital sensors and lower Hippocampus/Total grey matter ratio were significantly associated with a higher risk of progression to AD dementia (z = -5.45, p < 0.001 and z = -2.70, p = 0.007, respectively). Thus, MEG beta power provides information about the risk of future cognitive decline that is independent of that provided by neuroanatomical data. Higher education and lower MMSE showed additional but weaker prediction of increased probability of progression to AD dementia (z = 1.99, p = 0.05 and z = -1.95, p = 0.05, respectively).

**Figure 2:**
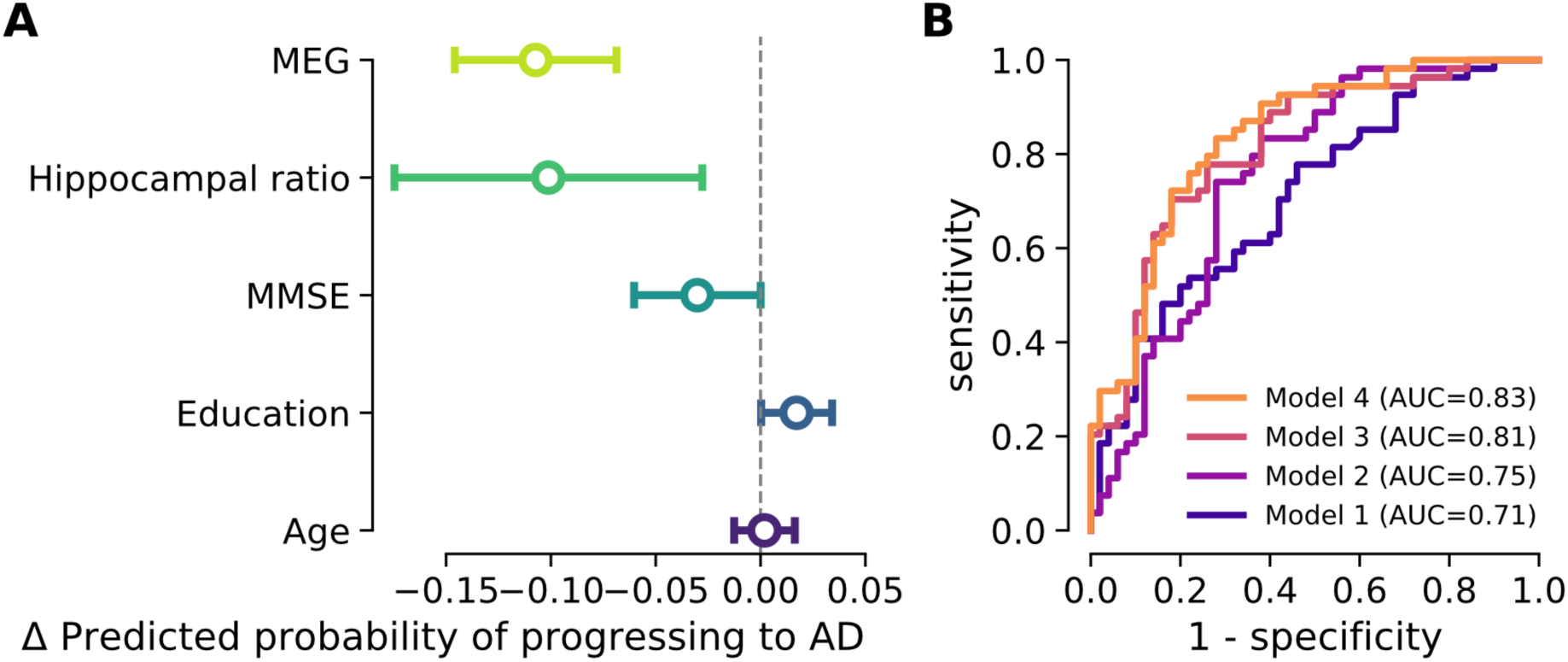
**(A)** Marginal effects of Hippocampus/Total grey matter ratio, MMSE, MEG 16-36Hz spectral power in parieto-occipital sensors, education and age on the probability of progression to AD dementia. Higher values of MEG 16-36Hz spectral power in left parieto-occipital sensors and higher Hippocampus/Total grey matter ratio were significantly associated with a reduced risk of progression to AD dementia. A higher level of education and lower MMSE had a trend towards increasing the probability of progression to AD dementia. (**B)** ROC curves of four logistic regression models to predict progression to AD dementia. Models including MEG perform better than other models.

We compared four logistic regression models for the risk of AD progression (**Figure 2B**) in terms of area under the curve (AUC). We first constructed a baseline with clinical and demographic variables only: Model 1 combined age, education and MMSE had a 0.71 AUC (65% sensitivity and 58% specificity). In a next step, we constructed an enhanced baseline including MRI: Model 2 combining age, education, MMSE and Hippocampus/Total grey matter ratio had a 0.75 AUC (72% sensitivity and 72% specificity). Then, we created a model that combined clinical and demographic variables with MEG to explore the added value of electrophysiology to clinical information: Model 3 combining age, education, MMSE and cluster MEG power better explained the data than Model 1 and 2, with a 0.81 AUC (76% sensitivity and 74% specificity). Thus, performances increased from Model 1 to Model 2 and performances improved markedly from Model 2 to Model 3. Finally, Model 4 combining age, education, MMSE, cluster MEG power and Hippocampus/Total grey matter ratio achieved slightly better results than Model 3 with a 0.83 AUC (78% sensitivity and 76% specificity).

### Analysis of robustness to spatial averaging

We performed a sensitivity analysis to assess the impact of spatial averaging over all sensors without using the cluster for multimodal analysis of AD progression risk. MEG 16-36 Hz average spectral power over all sensors (adjusted for MMSE), hippocampal ratio and MMSE remained additive in the logistic regression model (**Supplementary Figure 5A**). Models including MEG performed better than other models (**Supplementary Figure 5B**).

### Analysis of robustness to head position modelling

We performed an additional sensitivity analysis to assess the impact of head position alignment, using Maxfilter’s “trans – default” option **(Supplementary Figure 6 and 7).** Conclusions were unchanged for the averaged spectral power differences regardless of whether the power spectrum was adjusted for MMSE or not. The spatial pattern changed, but remained significant before adjusting for MMSE, though not when regressing out MMSE. MEG spectral power, hippocampal ratio and MMSE remained additive in the logistic regression model when using the cluster without MMSE correction (**Supplementary Table 1**).

## Discussion

In this study, we investigated the potential of MEG as a tool for predicting the progression from MCI to AD dementia in the BioFIND dataset. We analyzed data from 117 MCI patients, among whom 64 eventually developed AD dementia (AD progression), while 53 remained cognitively stable (stable MCI). Continuous spectral analysis of the power spectrum with Morlet Wavelets enabled us to avoid bias from pre-specified frequency bands and to define a common signal representation for various spectral measures, i.e., power, covariance, phase interactions and power envelopes. Additionally, Riemannian methods allowed fine-grained multivariate analysis of covariance matrices while reducing signal distortion. Our key findings revealed a significant reduction of MEG spectral power between 16Hz to 36Hz over parieto-occipital magnetometers in individuals who later progressed to AD dementia. This posterior 16Hz to 36Hz power reduction emerged as a robust predictor of future cognitive decline, even when considering conventional metrics like MMSE score and structural brain measures. Interestingly, adding MEG 16Hz to 36Hz power improved logistic regression models based on hippocampal/total grey matter ratio. Moreover, covariance matrices analyzed with Riemannian methods exhibited significant differences between groups, confirmed the differences in high frequencies (10-64Hz) and, in addition, uncovered low-frequency differences (< 4Hz). These findings highlight the potential of spectral power as a promising non-invasive electrophysiological biomarker to monitor AD progression, complementing classical diagnostic measures, including cognitive scores and structural MRI.

### Posterior MEG power (16Hz - 36 Hz) is reduced in AD progression

MCI patients who later progressed to AD dementia demonstrated reduced MEG power between 16 and 36 Hz, in the beta band, localized to the parieto-occipital sensors. This frequency-specific effect is in line with studies showing decreased power in alpha and beta bands in individuals with AD compared with healthy ageing, especially in the temporal and posterior/occipital brain regions (Babiloni et al., 2020; Cassani et al., 2018; Jelic et al., 2000; López-Sanz et al., 2019; Maestú & Fernández, 2020; Osipova et al., 2005; Stam et al., 2003). Claus et al. (1998) also reported that loss of beta band power was an independent predictor of an unfavourable prognosis in AD. Interestingly, a previous analysis of the BioFIND data (Vaghari, et al., 2022b) found that sensor covariance in the low gamma range (30-48Hz) was most informative, but this was for the classification of MCI versus controls (orthogonal to the convertor/stable MCI distinction used here).

Two principal hypotheses have been proposed to explain the ‘slowing’ of brain activity in AD, e.g. increased low-frequency and decreased high-frequency activity. The main hypothesis is based on the cholinergic deficit, as correlation between loss of cholinergic neurons and increased delta and theta power has been shown in patients with AD (Maestú & Fernández, 2020). Moreover, the administration of cholinergic antagonists in animal and human models have shown to induce delta and theta activity (Osipova et al., 2003). In our work, the AD progression group did not show increased delta nor theta power in the standard spectral analysis compared to the MCI stable group. However, our study revealed differences in the delta frequency band using Riemannian MANOVA, although without indicating the direction of these differences. The second hypothesis relies on AD being considered as a disconnection syndrome (Delbeuck et al., 2007). Cortico-thalamic disconnection in particular could play a role not only in the increased delta activity, but also in the decreased beta activity observed in AD, and as observed here, as suggested by Holschneider & Leuchter (1995). Further investigations would be needed to determine causative factors and provide a more comprehensive understanding of spectral power changes during AD progression.

### Posterior MEG power (16Hz - 36Hz) as a potential signature of cognitive function

Posterior MEG spectral power (16Hz - 36Hz) outperformed conventional metrics like the MMSE score and structural brain measures in predicting progression from MCI to AD dementia. Combining age, education, MMSE, and posterior MEG power (16Hz - 36Hz) achieved superior predictive performance than combining age, education and MMSE with hippocampal volume/total grey matter. These classification performances are in line with previous studies predicting conversion from MCI to AD dementia using EEG. Engedal et al. (2020) obtained effect sizes similar to those in our study with an AUC of 0.78, a sensitivity of 71%, a specificity of 69%, using a quantitative EEG Dementia Index and statistical pattern recognition method based on covariances. In our study, the fact that beta power emerged as a robust predictor of future cognitive decline is consistent with the study by Poil et al. (2013), which found that biomarkers sensitive to changes in the beta frequency (13–30 Hz) band were the most optimal for predicting conversion from MCI to AD, after assessing 177 candidate EEG biomarkers (Poil et al., 2013). These authors hypothesised that AD progression is associated with less stable beta frequency, possibly related to a less efficient working memory, given that beta oscillations are believed to maintain the current sensorimotor, cognitive state and attention (Engel & Fries, 2010; Gola et al., 2013). Huang et al. (2000) found that MCI patients who progressed to AD had a more anterior location of beta sources than stable MCI, and Baker et al. (2008) were able to classify MCI converters versus non-converters based on their EEG beta profile. On the other hand, increases in frontal beta power were observed in preclinical AD, and interpreted as compensatory mechanisms (Gaubert et al., 2019), suggesting a distinct mechanism, given the opposite direction of the effect and more anterior locus. Other EEG biomarkers found to be useful to predict decline from MCI to AD include higher alpha3/alpha2 frequency power ratio (Moretti, 2015), decreased posterior alpha power (Babiloni et al., 2020; Huang et al., 2000; Luckhaus et al., 2008), while (Rossini et al., 2006) described higher power values in the delta, theta, and alpha 1 bands, mainly over temporal and parietal areas in converters.

There are fewer studies using MEG to assess the progression from MCI to AD (Maestú & Fernández, 2020). A resting-state MEG study conducted by Fernández et al. (2006) identified higher delta power in a left parietal region as a reliable indicator of conversion within a 2-year period. López et al. (2014) found an increase in phase synchronization in the alpha band between the right anterior cingulate and temporo-occipital areas in AD converters. Our results are difficult to compare directly with these, because we only analysed power spectra in sensor-space, given our focus on more clinically-applicable MEG metrics (see Introduction).

### Low-frequency MEG activity (1Hz - 4Hz) uncovered by Riemannian analysis

Another important finding is that covariance matrices analyzed with Riemannian methods exhibited significant differences between groups across a wider range of frequencies than spectral power alone, particularly in the lower range of 1-4Hz, suggesting higher sensitivity of covariance versus power to identify future decliners. Riemannian tools have been shown to reduce signal distortion and are robust to field spread, potentially bypassing the need for source localization with a biophysical model (Chevallier et al., 2022; Sabbagh et al., 2020) while implying a logarithmic function similar to log power. These tools enabled us to capture signal changes in lower frequencies that were not consistently detectable based on pure sensor space power, which is systematically distorted by MEG field spread or EEG volume conduction. Importantly, prior work on age-prediction showed that the Riemannian embedding improved prediction performance compared to log power in sensor space (Sabbagh et al., 2020), reaching equivalence with source power analysis. However, adding Riemannian embeddings after source analysis did not improve performance (Sabbagh et al., 2020). We therefore propose our Riemannian signal as a potential surrogate for source power analysis, potentially improving diagnostic and prognostic biomarkers for neurodegenerative diseases, and therefore recommend them as features for future statistical modelling and machine learning analyses.

### MEG offers complementary functional information beyond structural damage

We found an additive effect of MEG and MRI in our study as both MEG beta power and Hippocampal Volume/Total Grey Matter Ratio were selected by AIC in the logistic regression model; i.e., they were able to bring unique independent information on the risk of cognitive decline. This aligns with prior research suggesting selective associations between hippocampal volume and specific EEG frequency bands (Meyer et al., 2017; Ruzich et al., 2019). Previous studies have demonstrated correlations between hippocampal volume and power in the alpha and theta bands, but not in the beta band (Babiloni et al., 2009; Grunwald et al., 2001, 2007; Moretti et al., 2007). Thus, we would cautiously suggest that the observed beta power effect is unrelated to hippocampal alterations. Nonetheless, more generally, our results reinforce claims that MEG (and EEG) offer complimentary information about MCI/AD beyond structural MRI measures alone (Vaghari et al., 2022b).

### Towards a “biomarker pyramid”: non-invasive electrophysiology as a key player in cognitive risk evaluation

In our study, we achieved improved modelling results by integrating age, education, MEG power, and a simple volumetric MRI metric (Hippocampal ratio). However, stronger effects would be required for definitive diagnosis, particularly when it implies decisions related to specific AD treatment in patients. This is in line with the proposition by Rossini et al. (2022) regarding a “biomarker pyramid” framework for cognitive risk evaluation, which foresees initial multimodal screening with widely accessible non-invasive methods (e.g. EEG, MRI, blood-based biomarkers, cognition) and follow-up or confirmation with expensive or invasive gold-standard approaches (PET, CSF). To realize this vision, it will be important to extend the use of EEG and/or MEG in clinical practice and research studies. Moreover, direct comparisons of EEG and MEG on the same participants would further support the goal of clinically validating MEG as a valuable additional clinical biomarker.

### Decreased phase- and amplitude interactions in the alpha band in AD progression

Despite finding that power spectra and sensor covariance were clearly able to distinguish converters from stable MCI, our results were less clear about the value of measures of functional coupling, namely dwPLI and envelope-correlation. While our results suggest decreased dwPLI and power envelope correlation in posterior brain regions within the alpha band in the AD progression group, this did not survive corrections for multiple comparisons, so needs replication. One advantage of our study was the use of a common representation based on Morlet wavelets (Bomatter et al., 2023; Hipp et al., 2012), which allowed us to compare dwPLI and power envelope correlation more directly with pure power spectra and sensor covariance. Importantly, it is well known that power is a principal confounder of connectivity, and that differences in signal-to-noise ratio can lead to spurious connectivity differences (Hawellek et al., 2022; Hipp et al., 2012). It is therefore interesting that, despite its weak effect size, our observation of reduced coupling between posterior regions occurred in the alpha range, rather than the beta range where we found the largest difference in power. This suggests that these potential changes in neural coupling associated with progression to AD dementia are not an artefact of differences in overall power.

Nonetheless, decreased alpha coupling contrasts with previous studies that have shown increased alpha synchronization in posterior regions in MCI converters, interpreted as compensatory mechanisms or neurotoxicity of amyloid load (Bajo et al., 2012; López et al., 2014). However, the functional connectivity pattern we describe resembles the one usually found in patients with AD dementia, e.g. reduced synchronization in alpha and beta frequency bands (Alonso et al., 2011; Babiloni et al., 2009, 2016; Gomez et al., 2012; Stam et al., 2003). One explanation could be that the MCI patients in our study were already more advanced in the disease course, as suggested by a mean MMSE of 25.5 in the AD progression group in our study, compared to 27.4 and 27.7 in the study by López et al. (2014) and Bajo et al. (2012) respectively. This would be consistent with the proposal of Pusil et al. (2019), that discrepancies between previous MEG/EEG comparisons of MCI versus controls, relative to comparisons of MCI versus AD dementia, reflect the possibility that functional connectivity follows an inverted-U shape as a function of disease progression, with increased (hyper)connectivity from healthy controls to MCI, followed by decreased (hypo)connectivity from MCI to AD dementia. Only very few studies have compared functional connectivity metrics in progressive and stable MCI patients (Bajo et al., 2012; López et al., 2014; Rossini et al., 2006), but this comparison should resemble more comparisons of MCI versus AD dementia (rather than healthy controls versus MCI), consistent at least with the decreases we found in coupling.

### Limitations and future directions

While our study provides valuable insights into the potential of non-invasive electrophysiology as a predictive tool for the progression from MCI to AD dementia, there are certain limitations that should be considered. Even if the clinical diagnosis of AD dementia was done by neurologists in specialized memory clinics, the lack of CSF or amyloid PET biomarkers introduces a risk of misdiagnosis approximating 30% (Dubois et al., 2023; Sabbagh et al., 2017). Incorporating CSF or PET biomarker assessments in future studies would refine the diagnostic specificity and enhance the reliability of the predictive models. Our study’s sample size is relatively large compared to other MEG or EEG studies on MCI progression (n=117), however it is still limited for developing prediction models. Moreover, in our study, patients showing AD progression presented lower MMSE scores at baseline compared to stable MCI, which raises the possibility that our results reflect a later stage of MCI at baseline, rather than solely capturing the difference between subsequent converters and non-converters. This highlights the importance of considering the temporal dynamics of disease progression in our interpretation, and of longitudinal MEG/EEG assessment (Lanskey et al., 2022).

Future studies should expand the scope of our work to characterize neurodegenerative diseases other than AD. Assessing these biomarkers in diseases like dementia with Lewy bodies (DLB), frontotemporal dementia (FTD), and Parkinson’s disease could provide a comprehensive understanding of their specificity and generalizability. Investigating how protein deposition alters electrophysiological patterns may uncover distinct pathways in various neurodegenerative disorders. Finally, our findings should be validated in EEG studies, going towards clinical application, as EEG is more widely available and cost-effective than MEG.

## Conclusion

This study combined MEG, anatomical MRI, cognitive and demographic information to address the potential of non-invasive electrophysiological biomarkers in monitoring the risk of progression from MCI to AD dementia. By exploring several spectral features computed using Morlet wavelets, we identified beta band power and covariance analyzed with Riemannian methods as promising biomarkers to help predict future cognitive decline. These findings hold promise for the development of screening strategies in large populations of MCI individuals and align with the emergence of new AD disease-modifying treatments.

## Data Availability

BioFIND is a controlled access repository and access is institutionally granted.

## Competing Interests

P.G., J.F.H. & D.E. are full-time employees of F. Hoffmann - La Roche Ltd.

## Author contributions

In alphabetical order.

**Conceptualization**: D.E., S.G.

**Data curation**: D.V., R.H.

**Formal analysis**: D.E., S.G.

**Investigation**: S.G.

**Methodology**: D.E., J.F.H., P.G., S.G.

**Project administration**: C.P., D.E, S.G.

**Software**: D.E., J.F.H.

**Supervision**: C.P., D.E.

**Validation**: D.E.

**Visualization**: D.E., S.G.

**Writing—original draft**: D.E., S.G

**Writing—review and editing**: C.P., D.E., D.V., J.F.H, ME.L., F.M., P.G., R.B., R.H., S.G.

## Supplementary figures

**Supplementary Figure 1:**
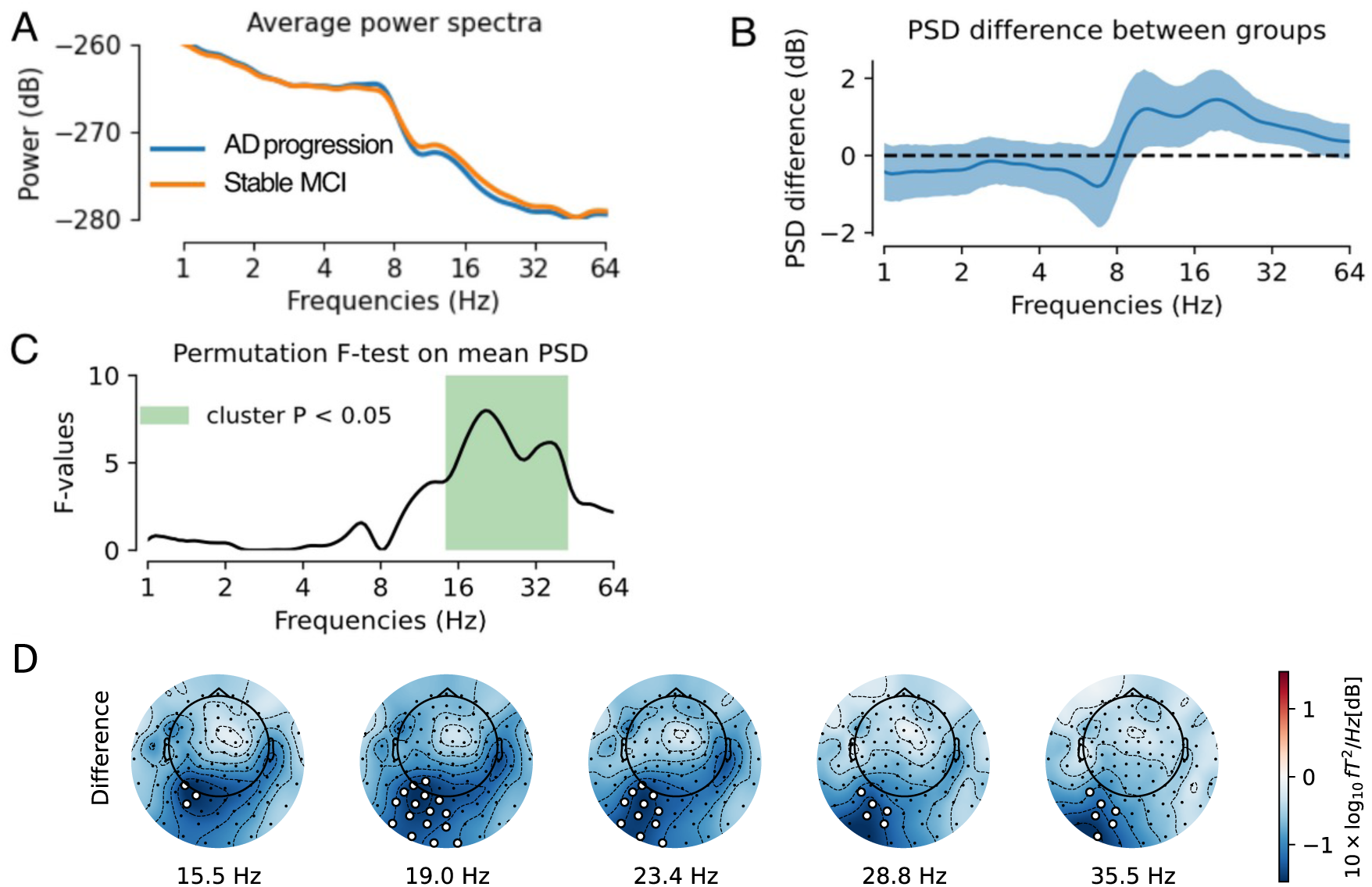
**(A)** Average power spectra over all sensors, adjusted on MMSE: AD progression group demonstrated reduced spectral power in frequencies ranging from 14 Hz to 44 Hz. **(B)** Mean spectral power difference between groups (blue line) and 95% confidence interval computed by bootstrap (blue shaded area). **(C)** Permutation F-test on mean spectral power, adjusted on MMSE, showing significant power difference between AD progression and stable MCI at frequencies ranging from 14.4 Hz to 43.7 Hz. **(D)** Topographical maps of spectral power difference between groups, adjusted on MMSE, showing reduced spectral power in left parieto-occipital region in AD progression group in frequencies from 15.5 Hz to 35.5 Hz.

**Supplementary Figure 2:**
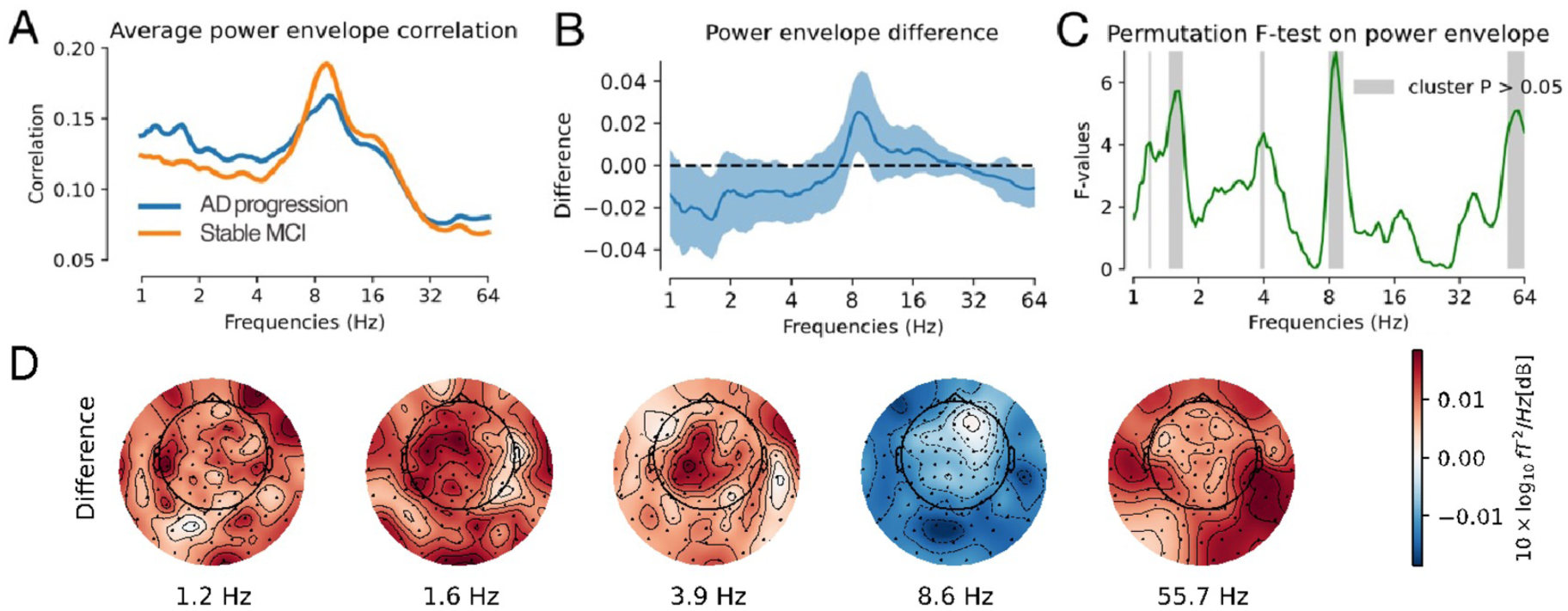
**(A)** Average power envelope correlation over all sensors: AD progression was visually associated with reduced power envelope correlation in alpha frequency band. **(B)** Power envelope correlation difference between groups (blue line) and 95% confidence interval computed by bootstrap (blue shaded area). The permutation test showed a p-value below 0.05 (uncorrected) for power envelope correlation at the following frequencies: 1.1Hz, 1.4Hz to 1.7Hz, 3.8Hz to 4Hz, 8Hz to 9.5Hz, 52Hz to 64Hz. **(C)** Permutation F-test with multiple-comparison correction across all frequencies for power envelope correlation, comparing AD progression group versus stable MCI, showing five clusters that did not reach significance, at the following frequencies: 1.1Hz, 1.4Hz to 1.7Hz, 3.8Hz to 4Hz, 8Hz to 9Hz, 53Hz to 64Hz. The cluster between 8Hz and 9Hz was the one with the smallest p-value (0.27). (**D)** Topographical maps of power envelope correlation differences between groups, shown at the five cluster frequencies. AD progression showed a pattern of decreased power envelope in alpha band in posterior regions, which did not reach statistical significance.

**Supplementary Figure 3:**
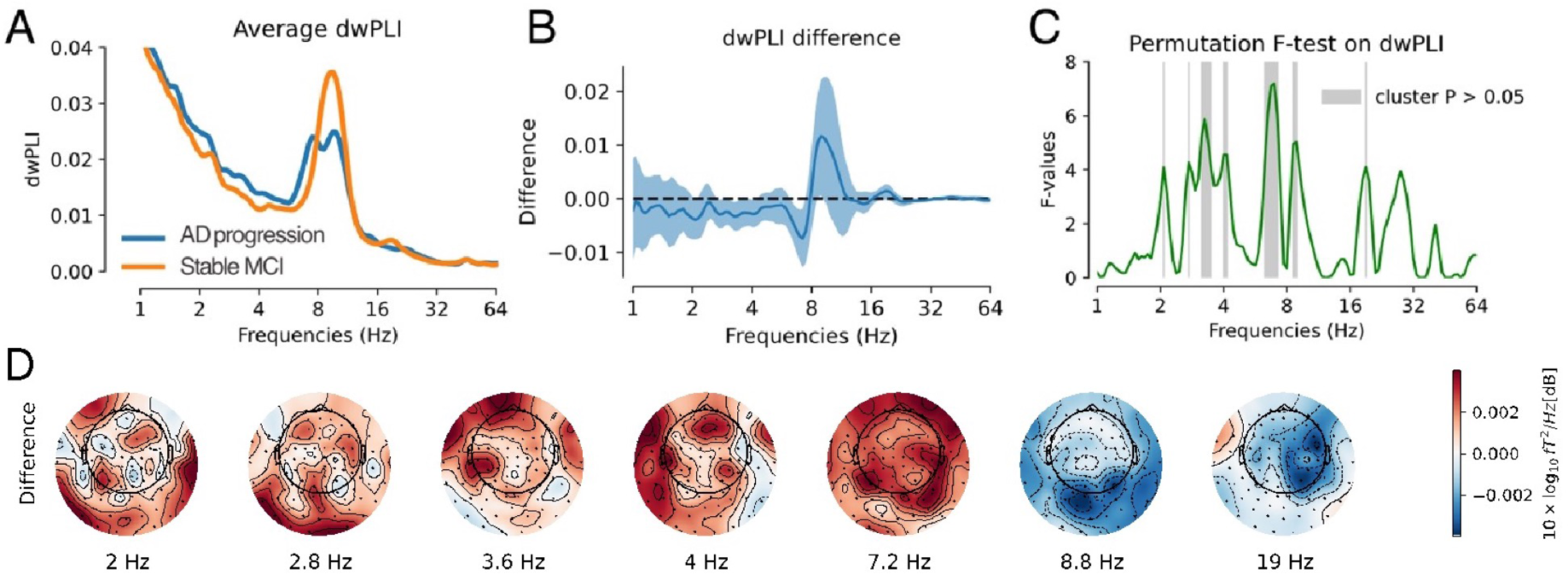
**(A)** Average dwPLI over all sensors: AD progression was visually associated with reduced dwPLI in alpha band. **(B)** dwPLI difference between groups (blue line) and 95% confidence interval computed by bootstrap (blue shaded area). The permutation test showed p-values below 0.05 (uncorrected) for dwPLI at the following frequencies: 2 Hz, 2.7 Hz, 2.8 Hz, 3.1-3.5 Hz, 3.9 Hz, 4.1Hz, 6.2-7.2Hz, 8.6-8.8Hz, 19-19.6Hz, 27.8Hz. (**C)** Permutation F-test with multiple-comparison correction across all frequencies for dwPLI, comparing AD progression group versus stable MCI, showing seven clusters that did not reach statistical significance, at the following frequencies: 2 Hz, 2.8 Hz, 3.1 Hz to 3.6 Hz, 4 Hz to 4.3 Hz, 6.3 Hz to 7.5 Hz, 8.6 Hz to 9.2 Hz and 19 Hz. The cluster between 6.3 Hz to 7.5 Hz was the one with the smallest p-value (0.21). (**D)** Topographical maps of dwPLI differences between groups, shown at the cluster frequencies. In posterior brain regions, AD progression showed a pattern of decreased dwPLI in alpha band and increased dwPLI in theta band, which did not reach statistical significance.

**Supplementary Figure 4:**
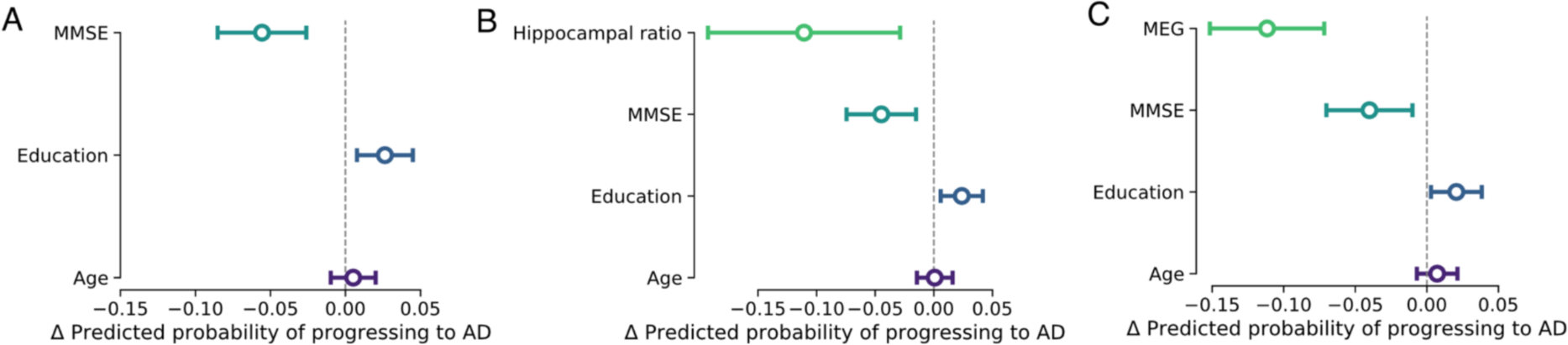
**(A)** Marginal effects of MMSE, education and age (Model 1) on the probability of progression to AD dementia. Lower MMSE and higher education were significantly associated with a higher risk of progression to AD dementia. (**B)** Marginal effects of Hippocampus/Total grey matter ratio, MMSE, education and age (Model 2) on the probability of progression to AD dementia. Lower hippocampal ratio, lower MMSE and higher education were significantly associated with a higher risk of progression to AD dementia. (**C)** Marginal effects of MEG 16-36Hz spectral power in parieto-occipital regions, MMSE, education and age (Model 3) on the probability of progression to AD dementia. Lower values of MEG 16-36Hz spectral power in left parieto-occipital region, lower MMSE and higher education were significantly associated with a higher risk of progression to AD dementia.

**Supplementary Figure 5:**
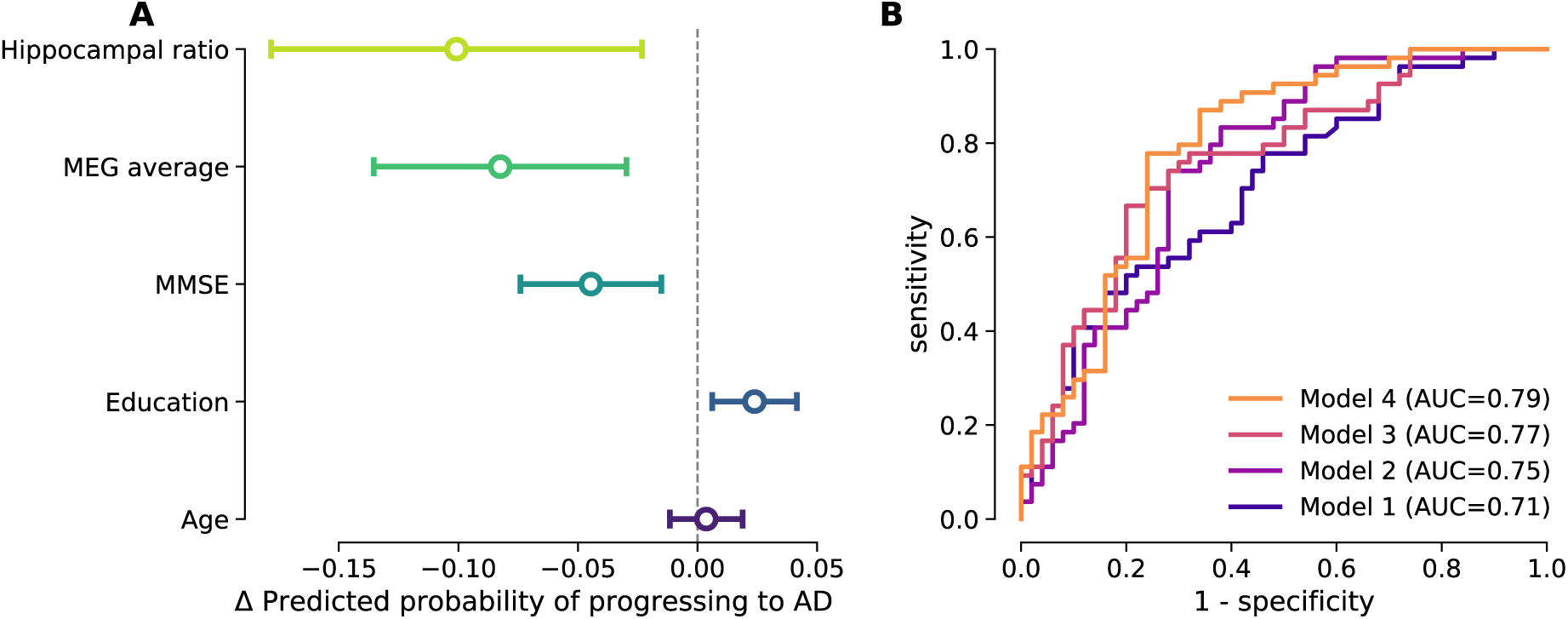
**(A)** Marginal effects of Hippocampus/Total grey matter ratio, MEG 16-36Hz average spectral power over all sensors (adjusted for MMSE), MMSE, education and age on the probability of progression to AD dementia. Higher values of MEG 16-36Hz average spectral power over all sensors, higher Hippocampus/Total grey matter ratio, higher MMSE and lower education were significantly associated with a reduced risk of progression to AD dementia. (**B)** ROC curves of four logistic regression models to predict progression to AD dementia. Model 1 combining age, education and MMSE had a 0.71 AUC (65% sensitivity and 58% specificity). Model 2 combining age, education, MMSE and Hippocampus/Total grey matter ratio had a 0.75 AUC (72% sensitivity and 72% specificity). Model 3 combining age, education, MMSE and average MEG 16-36 Hz power over all sensors had a 0.77 AUC (70% sensitivity and 72% specificity). Model 4 combining age, education, MMSE, cluster MEG power and Hippocampus/Total grey matter ratio achieved a 0.79 AUC (74% sensitivity and 76% specificity).

**Supplementary Figure 6:**
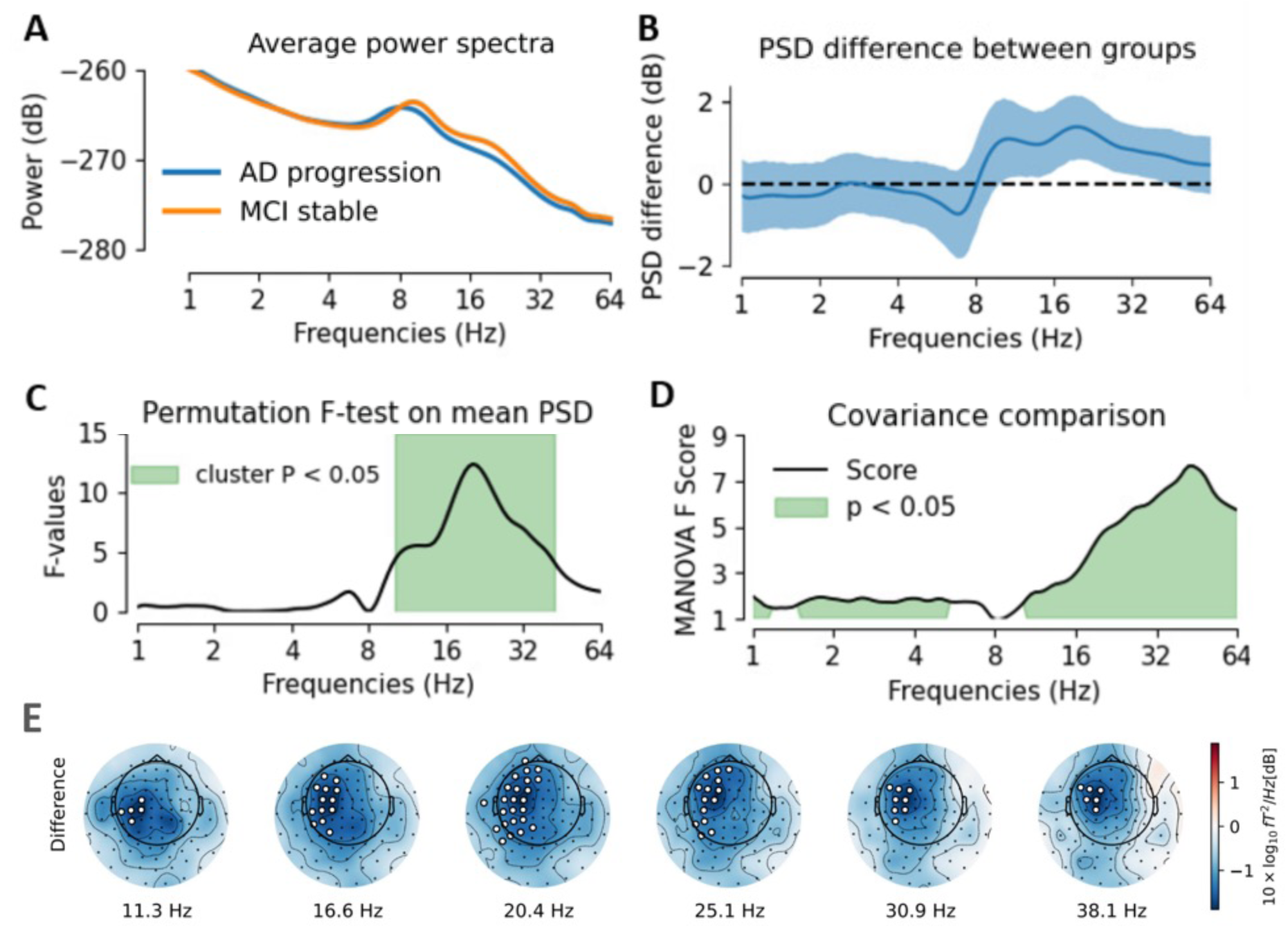
Sensitivity analysis results showing effect of head alignment. (**A)** Average power spectra over all sensors: AD progression was associated with reduced spectral power at baseline in frequencies ranging from 10 Hz to 44 Hz. (**B)** Mean spectral power difference between groups (blue line) and 95% confidence interval computed by bootstrap (blue shaded area). (**C)** Permutation F-test on mean spectral power, showing significant power difference between AD progression and stable MCI at frequencies ranging from 10 Hz to 44 Hz. (**D)** Comparison between groups based on frequency-specific covariances using distance Manova, showing significant differences between 1 Hz to 6 Hz and 10 Hz to 64 Hz. (**E)** Topographical maps of spectral power difference between groups, showing reduced spectral power in left fronto-temporo-parietal region in AD progression group in frequencies from 11 Hz to 38 Hz. The white dots indicate significant differences (p < 0.05)

**Supplementary Figure 7:**
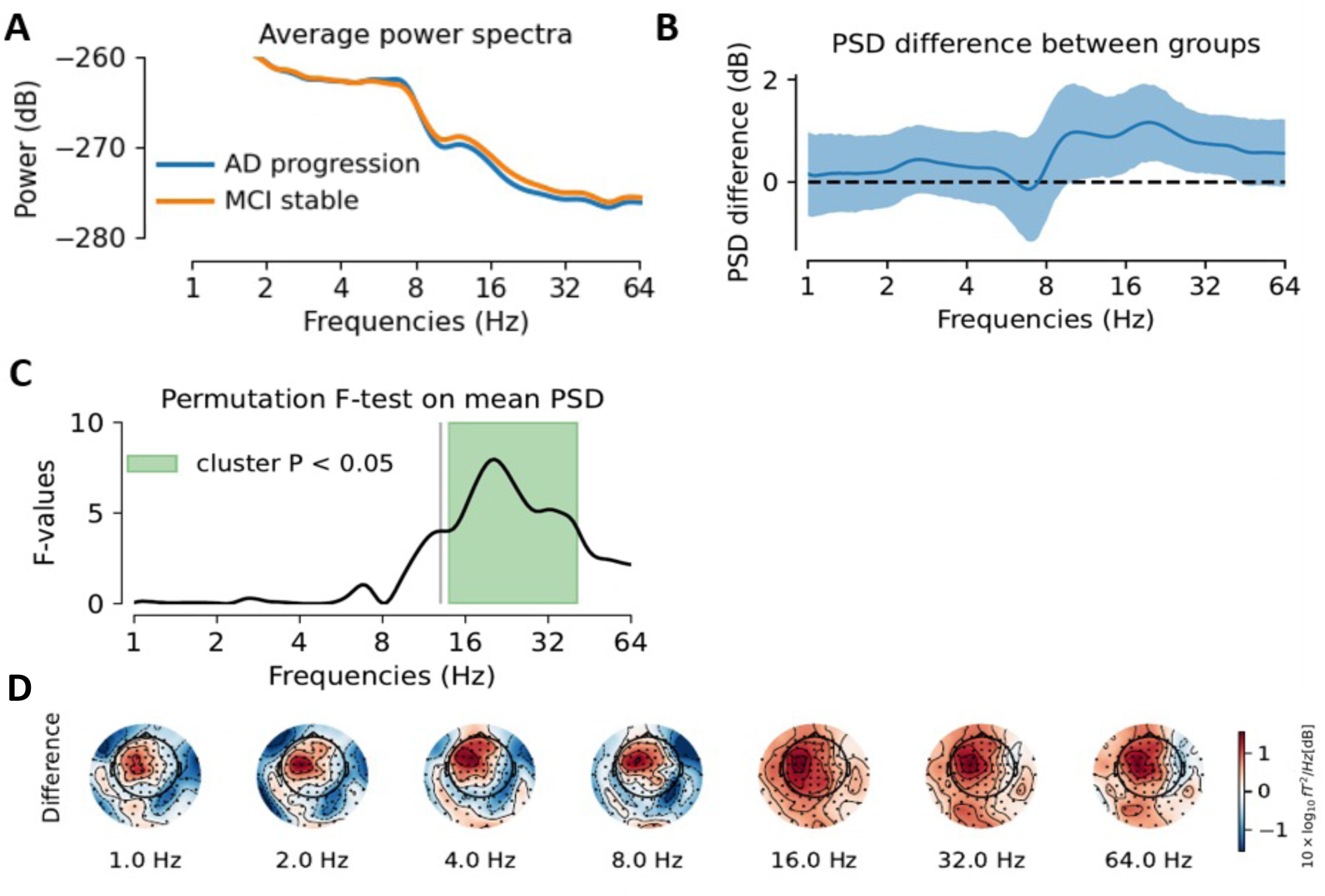
Sensitivity analysis results showing effect of head alignment, after adjustment on MMSE. **(A)** Average power spectra over all sensors, adjusted on MMSE: AD progression group demonstrated reduced spectral power in frequencies ranging from 14Hz to 42Hz. **(B)** Mean spectral power difference between groups (blue line) and 95% confidence interval computed by bootstrap (blue shaded area). **(C)** Permutation F-test on mean spectral power, adjusted on MMSE, showing significant power difference between AD progression and stable MCI at frequencies ranging from 14 Hz to 42 Hz. **(D)** Topographical maps of spectral power difference between groups showing no significant differences in spatial patterns after adjusting on MMSE.

**Supplementary Table 1:** Sensitivity analysis results showing effect of head alignment. Marginal effects of Hippocampus/Total grey matter ratio, MMSE, MEG 15-38Hz spectral power cluster in fronto-temporo-parietal regions, education and age on the probability of progression to AD dementia. Higher values of MEG 15-38Hz spectral power in the left fronto-temporo-parietal region and higher Hippocampus/Total grey matter ratio were significantly associated with a reduced risk of progression to AD dementia. A higher level of education and lower MMSE were associated with an increased probability of progression to AD dementia. *AME = Average Marginal Effect; SE = Standard Error, z = z-value, p = p-value, Lower = Lower Confidence Interval, Upper = Upper Confidence Interval*.

| Factor | AME | SE | z | p | Lower | Upper |
| --- | --- | --- | --- | --- | --- | --- |
| Age | 0.006 | 0.007 | 0.736 | 0.462 | -0.009 | 0.020 |
| Education | 0.021 | 0.009 | 2.375 | 0.018 | 0.004 | 0.039 |
| MEG (cluster) | -0.054 | 0.015 | -3.659 | 0.000 | -0.083 | -0.025 |
| MMSE | -0.034 | 0.015 | -2.295 | 0.022 | -0.063 | -0.005 |
| Hippocampal ratio | -0.106 | 0.039 | -2.738 | 0.006 | -0.182 | -0.030 |

## Notes

### Funding Statement

This work was supported by: EU JNPD (MR/P502017/1), MRC (SUAG/046 G101400) and Fondation pour la Recherche Médicale (grant FDM202106013579).

### Author Declarations

Local Ethics committee of the MRC Cognition & Brain Sciences Unit at the University of Cambridge, and of the Laboratory of Cognitive and Computational Neuroscience at the Centre for Biomedical Technology, Madrid, gave ethical approval for this work.

## References

Albert, M. S., DeKosky, S. T., Dickson, D., Dubois, B., Feldman, H. H., Fox, N. C., Gamst, A., Holtzman, D. M., Jagust, W. J., Petersen, R. C., Snyder, P. J., Carrillo, M. C., Thies, B., & Phelps, C. H. (2011). The diagnosis of mild cognitive impairment due to Alzheimer’s disease: Recommendations from the National Institute on Aging-Alzheimer’s Association workgroups on diagnostic guidelines for Alzheimer’s disease. Alzheimer’s & Dementia: The Journal of the Alzheimer’s Association, 7(3), 270–279. 10.1016/j.jalz.2011.03.008

Alonso, J. F., Poza, J., Mañanas, M. Á., Romero, S., Fernández, A., & Hornero, R. (2011). MEG Connectivity Analysis in Patients with Alzheimer’s Disease Using Cross Mutual Information and Spectral Coherence. Annals of Biomedical Engineering, 39(1), 524–536. 10.1007/s10439-010-0155-7

Alzheimer’s Association. (2023). 2023 Alzheimer’s disease facts and figures. Alzheimer’s & Dementia, 19(4), 1598–1695. 10.1002/alz.13016

Anderson, M. J. (2001). A new method for non-parametric multivariate analysis of variance. Austral Ecology, 26(1), 32–46. 10.1111/j.1442-9993.2001.01070.pp.x

Aydore, S., Pantazis, D., & Leahy, R. M. (2013). A note on the phase locking value and its properties. NeuroImage, 74, 231–244. 10.1016/j.neuroimage.2013.02.008

Babiloni, C., Binetti, G., Cassetta, E., Forno, G. D., Percio, C. D., Ferreri, F., Ferri, R., Frisoni, G., Hirata, K., Lanuzza, B., Miniussi, C., Moretti, D. V., Nobili, F., Rodriguez, G., Romani, G. L., Salinari, S., & Rossini, P. M. (2006). Sources of cortical rhythms change as a function of cognitive impairment in pathological aging: A multicenter study. Clinical Neurophysiology, 117(2), 252–268. 10.1016/j.clinph.2005.09.019

Babiloni, C., Frisoni, G., Pievani, M., Vecchio, F., Lizio, R., Buttiglione, M., Geroldi, C., Fracassi, C., Eusebi, F., & Ferri, R. (2009). Hippocampal volume and cortical sources of EEG alpha rhythms in mild cognitive impairment and Alzheimer disease. NeuroImage, 44(1), 123–135. 10.1016/j.neuroimage.2008.08.005

Babiloni, C., Lizio, R., Marzano, N., Capotosto, P., Soricelli, A., Triggiani, A. I., Cordone, S., Gesualdo, L., & Del Percio, C. (2016). Brain neural synchronization and functional coupling in Alzheimer’s disease as revealed by resting state EEG rhythms. International Journal of Psychophysiology, 103, 88–102. 10.1016/j.ijpsycho.2015.02.008

Babiloni, C., Lopez, S., Del Percio, C., Noce, G., Pascarelli, M. T., Lizio, R., Teipel, S. J., González-Escamilla, G., Bakardjian, H., George, N., Cavedo, E., Lista, S., Chiesa, P. A., Vergallo, A., Lemercier, P., Spinelli, G., Grothe, M. J., Potier, M.-C., Stocchi, F., … INSIGHT-preAD Study Group. (2020). Resting-state posterior alpha rhythms are abnormal in subjective memory complaint seniors with preclinical Alzheimer’s neuropathology and high education level: The INSIGHT-preAD study. Neurobiology of Aging, 90, 43–59. 10.1016/j.neurobiolaging.2020.01.012

Babiloni, C., Visser, P. J., Frisoni, G., De Deyn, P. P., Bresciani, L., Jelic, V., Nagels, G., Rodriguez, G., Rossini, P. M., Vecchio, F., Colombo, D., Verhey, F., Wahlund, L.-O., & Nobili, F. (2010). Cortical sources of resting EEG rhythms in mild cognitive impairment and subjective memory complaint. Neurobiology of Aging, 31(10), 1787–1798. 10.1016/j.neurobiolaging.2008.09.020

Bajo, R., Castellanos, N. P., Cuesta, P., Aurtenetxe, S., Garcia-Prieto, J., Gil-Gregorio, P., del-Pozo, F., & Maestu, F. (2012). Differential Patterns of Connectivity in Progressive Mild Cognitive Impairment. Brain Connectivity, 2(1), 21–24. 10.1089/brain.2011.0069

Baker, M., Akrofi, K., Schiffer, R., & Boyle, M. W. O. (2008). EEG Patterns in Mild Cognitive Impairment (MCI) Patients. The Open Neuroimaging Journal, 2, 52–55. 10.2174/1874440000802010052

Balart-Sánchez, S. A., Bittencourt-Villalpando, M., van der Naalt, J., & Maurits, N. M. (2021). Electroencephalography, Magnetoencephalography, and Cognitive Reserve: A Systematic Review. Archives of Clinical Neuropsychology, 36(7), 1374–1391. 10.1093/arclin/acaa132

Barachant, A., Barthélemy, Q., Gramfort, A., Jean-Rémi KING, Rodrigues, P. L. C., Dave, Olivetti, E., Goncharenko, V., Maxdolle, Berg, G. W. V., G. Reguig, Yamamoto, M. S., Artim436, Beasley, B., Bjäreholt, E., Clisson, P., Höchenberger, R., Jliersch, Sassenhagen, J., … Stonebig. (2023). pyRiemann/pyRiemann: V0.5 (v0.5) [Computer software]. Zenodo. 10.5281/ZENODO.8059038

Barachant, A., Bonnet, S., Congedo, M., & Jutten, C. (2012). Multiclass Brain–Computer Interface Classification by Riemannian Geometry. IEEE Transactions on Biomedical Engineering, 59(4), 920–928. 10.1109/TBME.2011.2172210

Benwell, C. S. Y., Davila-Pérez, P., Fried, P. J., Jones, R. N., Travison, T. G., Santarnecchi, E., Pascual-Leone, A., & Shafi, M. M. (2020). EEG spectral power abnormalities and their relationship with cognitive dysfunction in patients with Alzheimer’s disease and type 2 diabetes. Neurobiology of Aging, 85, 83–95. 10.1016/j.neurobiolaging.2019.10.004

Blinowska, K. J., Rakowski, F., Kaminski, M., De Vico Fallani, F., Del Percio, C., Lizio, R., & Babiloni, C. (2017). Functional and effective brain connectivity for discrimination between Alzheimer’s patients and healthy individuals: A study on resting state EEG rhythms. Clinical Neurophysiology, 128(4), 667–680. 10.1016/j.clinph.2016.10.002

Bomatter, P., Paillard, J., Garces, P., Hipp, J., & Engemann, D. (2023). Machine learning of brain-specific biomarkers from EEG (p. 2023.12.15.571864). bioRxiv. 10.1101/2023.12.15.571864

Bourguignon, M., Jousmäki, V., Dalal, S. S., Jerbi, K., & De Tiège, X. (2019). Coupling between human brain activity and body movements: Insights from non-invasive electromagnetic recordings. NeuroImage, 203, 116177. 10.1016/j.neuroimage.2019.116177

Brookes, M. J., Leggett, J., Rea, M., Hill, R. M., Holmes, N., Boto, E., & Bowtell, R. (2022). Magnetoencephalography with optically pumped magnetometers (OPM-MEG): The next generation of functional neuroimaging. Trends in Neurosciences, 45(8), 621–634. 10.1016/j.tins.2022.05.008

Brookes, M. J., Woolrich, M. W., & Barnes, G. R. (2012). Measuring functional connectivity in MEG: A multivariate approach insensitive to linear source leakage. NeuroImage, 63(2), 910–920. 10.1016/j.neuroimage.2012.03.048

Buzsáki, G., & Mizuseki, K. (2014). The log-dynamic brain: How skewed distributions affect network operations. Nature Reviews Neuroscience, 15(4), Article 4. 10.1038/nrn3687

Cassani, R., Estarellas, M., San-Martin, R., Fraga, F. J., & Falk, T. H. (2018). Systematic Review on Resting-State EEG for Alzheimer’s Disease Diagnosis and Progression Assessment. Disease Markers, 2018, 1–26. 10.1155/2018/5174815

Chevallier, S., Corsi, M.-C., Yger, F., & De Vico Fallani, F. (2022). Riemannian geometry for combining functional connectivity metrics and covariance in BCI. Software Impacts, 12, 100254. 10.1016/j.simpa.2022.100254

Claus, J. J., Ongerboer de Visser, B. W., Walstra, G. J., Hijdra, A., Verbeeten, B., & van Gool, W. A. (1998). Quantitative spectral electroencephalography in predicting survival in patients with early Alzheimer disease. Archives of Neurology, 55(8), 1105–1111. 10.1001/archneur.55.8.1105

Congedo, M., Barachant, A., & Bhatia, R. (2017). Riemannian geometry for EEG-based brain-computer interfaces; a primer and a review. Brain-Computer Interfaces, 4(3), 155–174. 10.1080/2326263X.2017.1297192

Dauwels, J., Vialatte, F., & Cichocki, A. (2010). Diagnosis of Alzheimers Disease from EEG Signals: Where Are We Standing? Current Alzheimer Research, 7(6), 487–505. 10.2174/156720510792231720

Delbeuck, X., Collette, F., & Van der Linden, M. (2007). Is Alzheimer’s disease a disconnection syndrome?: Evidence from a crossmodal audio-visual illusory experiment. Neuropsychologia, 45(14), 3315–3323. 10.1016/j.neuropsychologia.2007.05.001

Dubois, B., von Arnim, C. A. F., Burnie, N., Bozeat, S., & Cummings, J. (2023). Biomarkers in Alzheimer’s disease: Role in early and differential diagnosis and recognition of atypical variants. Alzheimer’s Research & Therapy, 15, 175. 10.1186/s13195-023-01314-6

Engedal, K., Barca, M. L., Høgh, P., Bo Andersen, B., Winther Dombernowsky, N., Naik, M., Gudmundsson, T. E., Øksengaard, A.-R., Wahlund, L.-O., & Snaedal, J. (2020). The Power of EEG to Predict Conversion from Mild Cognitive Impairment and Subjective Cognitive Decline to Dementia. Dementia and Geriatric Cognitive Disorders, 49(1), 38–47. 10.1159/000508392

Engel, A. K., & Fries, P. (2010). Beta-band oscillations—Signalling the status quo? Current Opinion in Neurobiology, 20(2), 156–165. 10.1016/j.conb.2010.02.015

Engemann, D. A., Kozynets, O., Sabbagh, D., Lemaître, G., Varoquaux, G., Liem, F., & Gramfort, A. (2020). Combining magnetoencephalography with magnetic resonance imaging enhances learning of surrogate-biomarkers. eLife, 9. 10.7554/eLife.54055

Ewers, M., Luan, Y., Frontzkowski, L., Neitzel, J., Rubinski, A., Dichgans, M., Hassenstab, J., Gordon, B. A., Chhatwal, J. P., Levin, J., Schofield, P., Benzinger, T. L. S., Morris, J. C., Goate, A., Karch, C. M., Fagan, A. M., McDade, E., Allegri, R., Berman, S., … Alzheimer’s Disease Neuroimaging Initiative and the Dominantly Inherited Alzheimer Network. (2021). Segregation of functional networks is associated with cognitive resilience in Alzheimer’s disease. Brain: A Journal of Neurology, 144(7), 2176–2185. 10.1093/brain/awab112

Fernández, A., Turrero, A., Zuluaga, P., Gil, P., Maestú, F., Campo, P., & Ortiz, T. (2006). Magnetoencephalographic Parietal δ Dipole Density in Mild Cognitive Impairment: Preliminary Results of a Method to Estimate the Risk of Developing Alzheimer Disease. Archives of Neurology, 63(3), 427–430. 10.1001/archneur.63.3.427

Fischl, B. (2012). FreeSurfer. NeuroImage, 62(2), 774–781. 10.1016/j.neuroimage.2012.01.021

Frey, J. N., Mainy, N., Lachaux, J.-P., Müller, N., Bertrand, O., & Weisz, N. (2014). Selective Modulation of Auditory Cortical Alpha Activity in an Audiovisual Spatial Attention Task. The Journal of Neuroscience, 34(19), 6634–6639. 10.1523/JNEUROSCI.4813-13.2014

Frohlich, J., Miller, M. T., Bird, L. M., Garces, P., Purtell, H., Hoener, M. C., Philpot, B. D., Sidorov, M. S., Tan, W.-H., Hernandez, M.-C., Rotenberg, A., Jeste, S. S., Krishnan, M., Khwaja, O., & Hipp, J. F. (2019). Electrophysiological Phenotype in Angelman Syndrome Differs Between Genotypes. Biological Psychiatry, 85(9), 752–759. 10.1016/j.biopsych.2019.01.008

Garcés, P., López-Sanz, D., Maestú, F., & Pereda, E. (2017). Choice of Magnetometers and Gradiometers after Signal Space Separation. Sensors, 17, 2926. 10.3390/s17122926

Garcés, P., Vicente, R., Wibral, M., Pineda-Pardo, J. Á., López, M. E., Aurtenetxe, S., Marcos, A., de Andrés, M. E., Yus, M., Sancho, M., Maestú, F., & Fernández, A. (2013). Brain-wide slowing of spontaneous alpha rhythms in mild cognitive impairment. Frontiers in Aging Neuroscience, 5, 100. 10.3389/fnagi.2013.00100

Gaubert, S., Raimondo, F., Houot, M., Corsi, M.-C., Naccache, L., Diego Sitt, J., Hermann, B., Oudiette, D., Gagliardi, G., Habert, M.-O., Dubois, B., De Vico Fallani, F., Bakardjian, H., & Epelbaum, S. (2019). EEG evidence of compensatory mechanisms in preclinical Alzheimer’s disease. Brain, 142(7), 2096–2112. 10.1093/brain/awz150

Gola, M., Magnuski, M., Szumska, I., & Wróbel, A. (2013). EEG beta band activity is related to attention and attentional deficits in the visual performance of elderly subjects. International Journal of Psychophysiology, 89(3), 334–341. 10.1016/j.ijpsycho.2013.05.007

Gomez, C., Martinez-Zarzuela, M., Poza, J., Diaz-Pernas, F. J., Fernandez, A., & Hornero, R. (2012). Synchrony analysis of spontaneous MEG activity in Alzheimer’s disease patients. 2012 Annual International Conference of the IEEE Engineering in Medicine and Biology Society, 6188–6191. 10.1109/EMBC.2012.6347407

Gramfort, A., Luessi, M., Larson, E., Engemann, D., Strohmeier, D., Brodbeck, C., Goj, R., Jas, M., Brooks, T., Parkkonen, L., & Hämäläinen, M. (2013). MEG and EEG data analysis with MNE-Python. Frontiers in Neuroscience, 7. https://www.frontiersin.org/articles/10.3389/fnins.2013.00267

Grunwald, M., Busse, F., Hensel, A., Kruggel, F., Riedel-Heller, S., Wolf, H., Arendt, T., & Gertz, H. J. (2001). Correlation between cortical theta activity and hippocampal volumes in health, mild cognitive impairment, and mild dementia. Journal of Clinical Neurophysiology: Official Publication of the American Electroencephalographic Society, 18(2), 178–184.

Grunwald, M., Hensel, A., Wolf, H., Weiss, T., & Gertz, H.-J. (2007). Does the hippocampal atrophy correlate with the cortical theta power in elderly subjects with a range of cognitive impairment? Journal of Clinical Neurophysiology: Official Publication of the American Electroencephalographic Society, 24(1), 22–26. 10.1097/WNP.0b013e31802ed5b2

Hampel, H., & Lista, S. (2016). Dementia: The rising global tide of cognitive impairment. Nature Reviews. Neurology, 12(3), 131–132. 10.1038/nrneurol.2015.250

Hawellek, D. J., Garces, P., Meghdadi, A. H., Waninger, S., Smith, A., Manchester, M., Schobel, S. A., & Hipp, J. F. (2022). Changes in brain activity with tominersen in early-manifest Huntington’s disease. Brain Communications, 4(3), fcac149. 10.1093/braincomms/fcac149

Hipp, J. F., Hawellek, D. J., Corbetta, M., Siegel, M., & Engel, A. K. (2012). Large-scale cortical correlation structure of spontaneous oscillatory activity. Nature Neuroscience, 15(6), Article 6. 10.1038/nn.3101

Hipp, J. F., Knoflach, F., Comley, R., Ballard, T. M., Honer, M., Trube, G., Gasser, R., Prinssen, E., Wallace, T. L., Rothfuss, A., Knust, H., Lennon-Chrimes, S., Derks, M., Bentley, D., Squassante, L., Nave, S., Nöldeke, J., Wandel, C., Thomas, A. W., & Hernandez, M.-C. (2021). Basmisanil, a highly selective GABAA-α5 negative allosteric modulator: Preclinical pharmacology and demonstration of functional target engagement in man. Scientific Reports, 11(1), 7700. 10.1038/s41598-021-87307-7

Holschneider, D. P., & Leuchter, A. F. (1995). Beta activity in aging and dementia. Brain Topography, 8(2), 169–180. 10.1007/BF01199780

Hu, X., Meier, M., & Pruessner, J. (2023). Challenges and opportunities of diagnostic markers of Alzheimer’s disease based on structural magnetic resonance imaging. Brain and Behavior, 13(3), e2925. 10.1002/brb3.2925

Huang, C., Wahlund, L., Dierks, T., Julin, P., Winblad, B., & Jelic, V. (2000). Discrimination of Alzheimer’s disease and mild cognitive impairment by equivalent EEG sources: A cross-sectional and longitudinal study. Clinical Neurophysiology: Official Journal of the International Federation of Clinical Neurophysiology, 111(11), 1961–1967. 10.1016/s1388-2457(00)00454-5

Jack, C. R., Bennett, D. A., Blennow, K., Carrillo, M. C., Dunn, B., Haeberlein, S. B., Holtzman, D. M., Jagust, W., Jessen, F., Karlawish, J., Liu, E., Molinuevo, J. L., Montine, T., Phelps, C., Rankin, K. P., Rowe, C. C., Scheltens, P., Siemers, E., Snyder, H. M., … Contributors. (2018). NIA-AA Research Framework: Toward a biological definition of Alzheimer’s disease. Alzheimer’s & Dementia: The Journal of the Alzheimer’s Association, 14(4), 535–562. 10.1016/j.jalz.2018.02.018

Janelidze, S., Bali, D., Ashton, N. J., Barthélemy, N. R., Vanbrabant, J., Stoops, E., Vanmechelen, E., He, Y., Dolado, A. O., Triana-Baltzer, G., Pontecorvo, M. J., Zetterberg, H., Kolb, H., Vandijck, M., Blennow, K., Bateman, R. J., & Hansson, O. (2023). Head-to-head comparison of 10 plasma phospho-tau assays in prodromal Alzheimer’s disease. Brain: A Journal of Neurology, 146(4), 1592–1601. 10.1093/brain/awac333

Jas, M., Engemann, D. A., Bekhti, Y., Raimondo, F., & Gramfort, A. (2017). Autoreject: Automated artifact rejection for MEG and EEG data. NeuroImage, 159, 417–429. 10.1016/j.neuroimage.2017.06.030

Jelic, V., Johansson, S. E., Almkvist, O., Shigeta, M., Julin, P., Nordberg, A., Winblad, B., & Wahlund, L. O. (2000). Quantitative electroencephalography in mild cognitive impairment: Longitudinal changes and possible prediction of Alzheimer’s disease. Neurobiology of Aging, 21(4), 533–540.

Jeong, J. (2004). EEG dynamics in patients with Alzheimer’s disease. Clinical Neurophysiology, 115(7), 1490–1505. 10.1016/j.clinph.2004.01.001

Jobert, M., & Wilson, F. J. (2015). Advanced Analysis of Pharmaco-EEG Data in Humans. Neuropsychobiology, 72(3–4), 165–177. 10.1159/000431096

King, J.-R., & Dehaene, S. (2014). Characterizing the dynamics of mental representations: The temporal generalization method. Trends in Cognitive Sciences, 18(4), 203–210. 10.1016/j.tics.2014.01.002

Lanskey, J. H., Kocagoncu, E., Quinn, A. J., Cheng, Y.-J., Karadag, M., Pitt, J., Lowe, S., Perkinton, M., Raymont, V., Singh, K. D., Woolrich, M., Nobre, A. C., Henson, R. N., & Rowe, J. B. (2022). New Therapeutics in Alzheimer’s Disease Longitudinal Cohort study (NTAD): Study protocol. BMJ Open, 12(12), e055135. 10.1136/bmjopen-2021-055135

Lee, D. H., Seo, S. W., Roh, J. H., Oh, M., Oh, J. S., Oh, S. J., Kim, J. S., & Jeong, Y. (2022). Effects of Cognitive Reserve in Alzheimer’s Disease and Cognitively Unimpaired Individuals. Frontiers in Aging Neuroscience, 13. https://www.frontiersin.org/articles/10.3389/fnagi.2021.784054

Leeper, T. J. (2017). Interpreting Regression Results using Average Marginal Effects with R ’ s margins. https://www.semanticscholar.org/paper/Interpreting-Regression-Results-using-Average-with-Leeper/9615c76bd5d81f7ebbbdac9714619863dc3a2337

Lehtelä, L., Salmelin, R., & Hari, R. (1997). Evidence for reactive magnetic 10-Hz rhythm in the human auditory cortex. Neuroscience Letters, 222(2), 111–114. 10.1016/S0304-3940(97)13361-4

Li, R.-X., Ma, Y.-H., Tan, L., & Yu, J.-T. (2022). Prospective biomarkers of Alzheimer’s disease: A systematic review and meta-analysis. Ageing Research Reviews, 81, 101699. 10.1016/j.arr.2022.101699

López, M. E., Bruña, R., Aurtenetxe, S., Pineda-Pardo, J. Á., Marcos, A., Arrazola, J., Reinoso, A. I., Montejo, P., Bajo, R., & Maestú, F. (2014). Alpha-band hypersynchronization in progressive mild cognitive impairment: A magnetoencephalography study. The Journal of Neuroscience: The Official Journal of the Society for Neuroscience, 34(44), 14551–14559. 10.1523/JNEUROSCI.0964-14.2014

López-Sanz, D., Bruña, R., de Frutos-Lucas, J., & Maestú, F. (2019). 1b— Magnetoencephalography applied to the study of Alzheimer’s disease. In J. T. Becker & A. D. Cohen (Eds.), Progress in Molecular Biology and Translational Science (Vol. 165, pp. 25–61). Academic Press. 10.1016/bs.pmbts.2019.04.007

Luckhaus, C., Grass-Kapanke, B., Blaeser, I., Ihl, R., Supprian, T., Winterer, G., Zielasek, J., & Brinkmeyer, J. (2008). Quantitative EEG in progressing vs stable mild cognitive impairment (MCI): Results of a 1-year follow-up study. International Journal of Geriatric Psychiatry, 23(11), 1148–1155. 10.1002/gps.2042

Maestú, F., & Fernández, A. (2020). Role of Magnetoencephalography in the Early Stages of Alzheimer Disease. Neuroimaging Clinics of North America, 30(2), 217–227. 10.1016/j.nic.2020.01.003

Marizzoni, M., Ferrari, C., Jovicich, J., Albani, D., Babiloni, C., Cavaliere, L., Didic, M., Forloni, G., Galluzzi, S., Hoffmann, K.-T., Molinuevo, J. L., Nobili, F., Parnetti, L., Payoux, P., Ribaldi, F., Rossini, P. M., Schönknecht, P., Salvatore, M., Soricelli, A., … PharmaCog Consortium. (2019). Predicting and Tracking Short Term Disease Progression in Amnestic Mild Cognitive Impairment Patients with Prodromal Alzheimer’s Disease: Structural Brain Biomarkers. Journal of Alzheimer’s Disease: JAD, 69(1), 3–14. 10.3233/JAD-180152

McKhann, G. M., Knopman, D. S., Chertkow, H., Hyman, B. T., Jack, C. R., Kawas, C. H., Klunk, W. E., Koroshetz, W. J., Manly, J. J., Mayeux, R., Mohs, R. C., Morris, J. C., Rossor, M. N., Scheltens, P., Carrillo, M. C., Thies, B., Weintraub, S., & Phelps, C. H. (2011). The diagnosis of dementia due to Alzheimer’s disease: Recommendations from the National Institute on Aging-Alzheimer’s Association workgroups on diagnostic guidelines for Alzheimer’s disease. Alzheimer’s & Dementia, 7(3), 263–269. 10.1016/j.jalz.2011.03.005

Meyer, S. S., Rossiter, H., Brookes, M. J., Woolrich, M. W., Bestmann, S., & Barnes, G. R. (2017). Using generative models to make probabilistic statements about hippocampal engagement in MEG. NeuroImage, 149, 468–482. 10.1016/j.neuroimage.2017.01.029

Moretti, D. V. (2015). Conversion of mild cognitive impairment patients in Alzheimer’s disease: Prognostic value of Alpha3/Alpha2 electroencephalographic rhythms power ratio. Alzheimer’s Research & Therapy, 7, 80. 10.1186/s13195-015-0162-x

Moretti, D. V., Miniussi, C., Frisoni, G. B., Geroldi, C., Zanetti, O., Binetti, G., & Rossini, P. M. (2007). Hippocampal atrophy and EEG markers in subjects with mild cognitive impairment. Clinical Neurophysiology: Official Journal of the International Federation of Clinical Neurophysiology, 118(12), 2716–2729. 10.1016/j.clinph.2007.09.059

Musaeus, C. S., Nielsen, M. S., Musaeus, J. S., & Høgh, P. (2020). Electroencephalographic Cross-Frequency Coupling as a Sign of Disease Progression in Patients With Mild Cognitive Impairment: A Pilot Study. Frontiers in Neuroscience, 14, 790. 10.3389/fnins.2020.00790

Osipova, D., Ahveninen, J., Jensen, O., Ylikoski, A., & Pekkonen, E. (2005). Altered generation of spontaneous oscillations in Alzheimer’s disease. NeuroImage, 27(4), 835–841. 10.1016/j.neuroimage.2005.05.011

Osipova, D., Ahveninen, J., Kaakkola, S., Jääskeläinen, I. P., Huttunen, J., & Pekkonen, E. (2003). Effects of scopolamine on MEG spectral power and coherence in elderly subjects. Clinical Neurophysiology: Official Journal of the International Federation of Clinical Neurophysiology, 114(10), 1902–1907. 10.1016/s1388-2457(03)00165-2

Petersen, R. C., Caracciolo, B., Brayne, C., Gauthier, S., Jelic, V., & Fratiglioni, L. (2014). Mild cognitive impairment: A concept in evolution. Journal of Internal Medicine, 275(3), 214–228. 10.1111/joim.12190

Poil, S.-S., de Haan, W., van der Flier, W. M., Mansvelder, H. D., Scheltens, P., & Linkenkaer-Hansen, K. (2013). Integrative EEG biomarkers predict progression to Alzheimer’s disease at the MCI stage. Frontiers in Aging Neuroscience, 5. 10.3389/fnagi.2013.00058

Pusil, S., López, M. E., Cuesta, P., Bruña, R., Pereda, E., & Maestú, F. (2019). Hypersynchronization in mild cognitive impairment: The ‘X’ model. Brain, 142(12), 3936– 3950. 10.1093/brain/awz320

Rathore, S., Habes, M., Iftikhar, M. A., Shacklett, A., & Davatzikos, C. (2017). A review on neuroimaging-based classification studies and associated feature extraction methods for Alzheimer’s disease and its prodromal stages. NeuroImage, 155, 530–548. 10.1016/j.neuroimage.2017.03.057

Rossini, P. M., Del Percio, C., Pasqualetti, P., Cassetta, E., Binetti, G., Dal Forno, G., Ferreri, F., Frisoni, G., Chiovenda, P., Miniussi, C., Parisi, L., Tombini, M., Vecchio, F., & Babiloni, C. (2006). Conversion from mild cognitive impairment to Alzheimer’s disease is predicted by sources and coherence of brain electroencephalography rhythms. Neuroscience, 143(3), 793–803. 10.1016/j.neuroscience.2006.08.049

Rossini, P. M., Miraglia, F., & Vecchio, F. (2022). Early dementia diagnosis, MCI-to-dementia risk prediction, and the role of machine learning methods for feature extraction from integrated biomarkers, in particular for EEG signal analysis. Alzheimer’s & Dementia: The Journal of the Alzheimer’s Association, 18(12), 2699–2706. 10.1002/alz.12645

Ruzich, E., Crespo-García, M., Dalal, S. S., & Schneiderman, J. F. (2019). Characterizing hippocampal dynamics with MEG: A systematic review and evidence-based guidelines. Human Brain Mapping, 40(4), 1353–1375. 10.1002/hbm.24445

Sabbagh, D., Ablin, P., Varoquaux, G., Gramfort, A., & Engemann, D. A. (2019). Manifold-regression to predict from MEG/EEG brain signals without source modeling. Advances in Neural Information Processing Systems, 32. https://proceedings.neurips.cc/paper_files/paper/2019/hash/d464b5ac99e74462f321c06ccacc4bff-Abstract.html

Sabbagh, D., Ablin, P., Varoquaux, G., Gramfort, A., & Engemann, D. A. (2020). Predictive regression modeling with MEG/EEG: From source power to signals and cognitive states. NeuroImage, 222, 116893. 10.1016/j.neuroimage.2020.116893

Sabbagh, M. N., Lue, L.-F., Fayard, D., & Shi, J. (2017). Increasing Precision of Clinical Diagnosis of Alzheimer’s Disease Using a Combined Algorithm Incorporating Clinical and Novel Biomarker Data. Neurology and Therapy, 6(Suppl 1), 83–95. 10.1007/s40120-017-0069-5

Schnitzler, A., & Gross, J. (2005). Normal and pathological oscillatory communication in the brain. Nature Reviews. Neuroscience, 6(4), 285–296. 10.1038/nrn1650

Siegel, M., Donner, T. H., & Engel, A. K. (2012). Spectral fingerprints of large-scale neuronal interactions. Nature Reviews. Neuroscience, 13(2), 121–134. 10.1038/nrn3137

Sims, J. R., Zimmer, J. A., Evans, C. D., Lu, M., Ardayfio, P., Sparks, J., Wessels, A. M., Shcherbinin, S., Wang, H., Monkul Nery, E. S., Collins, E. C., Solomon, P., Salloway, S., Apostolova, L. G., Hansson, O., Ritchie, C., Brooks, D. A., Mintun, M., Skovronsky, D. M., … Zboch, M. (2023). Donanemab in Early Symptomatic Alzheimer Disease: The TRAILBLAZER-ALZ 2 Randomized Clinical Trial. JAMA. 10.1001/jama.2023.13239

Smith, S. M., & Nichols, T. E. (2009). Threshold-free cluster enhancement: Addressing problems of smoothing, threshold dependence and localisation in cluster inference. NeuroImage, 44(1), 83–98. 10.1016/j.neuroimage.2008.03.061

Stam, C. J. (2010). Use of magnetoencephalography (MEG) to study functional brain networks in neurodegenerative disorders. Journal of the Neurological Sciences, 289(1–2), 128–134. 10.1016/j.jns.2009.08.028

Stam, C. J., van der Made, Y., Pijnenburg, Y. A. L., & Scheltens, P. (2003). EEG synchronization in mild cognitive impairment and Alzheimer’s disease. Acta Neurologica Scandinavica, 108(2), 90–96. 10.1034/j.1600-0404.2003.02067.x

Stern, Y. (2009). Cognitive Reserve. Neuropsychologia, 47(10), 2015. 10.1016/j.neuropsychologia.2009.03.004

Stokes, M. G., Wolff, M. J., & Spaak, E. (2015). Decoding Rich Spatial Information with High Temporal Resolution. Trends in Cognitive Sciences, 19(11), 636–638. 10.1016/j.tics.2015.08.016

Thijssen, E. H., La Joie, R., Strom, A., Fonseca, C., Iaccarino, L., Wolf, A., Spina, S., Allen, I. E., Cobigo, Y., Heuer, H., VandeVrede, L., Proctor, N. K., Lago, A. L., Baker, S., Sivasankaran, R., Kieloch, A., Kinhikar, A., Yu, L., Valentin, M.-A., … Advancing Research and Treatment for Frontotemporal Lobar Degeneration investigators. (2021). Plasma phosphorylated tau 217 and phosphorylated tau 181 as biomarkers in Alzheimer’s disease and frontotemporal lobar degeneration: A retrospective diagnostic performance study. The Lancet. Neurology, 20(9), 739–752. 10.1016/S1474-4422(21)00214-3

Uusitalo, M. A., & Ilmoniemi, R. J. (1997). Signal-space projection method for separating MEG or EEG into components. Medical and Biological Engineering and Computing, 35(2), 135–140. 10.1007/BF02534144

Vaghari, D., Bruna, R., Hughes, L. E., Nesbitt, D., Tibon, R., Rowe, J. B., Maestu, F., & Henson, R. N. (2022a). A multi-site, multi-participant magnetoencephalography resting-state dataset to study dementia: The BioFIND dataset. NeuroImage, 258, 119344. 10.1016/j.neuroimage.2022.119344

Vaghari, D., Kabir, E., & Henson, R. N. (2022b). Late combination shows that MEG adds to MRI in classifying MCI versus controls. NeuroImage, 252, 119054. 10.1016/j.neuroimage.2022.119054

Van Dyck, C. H., Swanson, C. J., Aisen, P., Bateman, R. J., Chen, C., Gee, M., Kanekiyo, M., Li, D., Reyderman, L., Cohen, S., Froelich, L., Katayama, S., Sabbagh, M., Vellas, B., Watson, D., Dhadda, S., Irizarry, M., Kramer, L. D., & Iwatsubo, T. (2023). Lecanemab in Early Alzheimer’s Disease. New England Journal of Medicine, 388(1), 9–21. 10.1056/NEJMoa2212948

Vinck, M., Oostenveld, R., van Wingerden, M., Battaglia, F., & Pennartz, C. M. A. (2011). An improved index of phase-synchronization for electrophysiological data in the presence of volume-conduction, noise and sample-size bias. NeuroImage, 55(4), 1548–1565. 10.1016/j.neuroimage.2011.01.055

Virtanen, P., Gommers, R., Oliphant, T. E., Haberland, M., Reddy, T., Cournapeau, D., Burovski, E., Peterson, P., Weckesser, W., Bright, J., van der Walt, S. J., Brett, M., Wilson, J., Millman, K. J., Mayorov, N., Nelson, A. R. J., Jones, E., Kern, R., Larson, E., … van Mulbregt, P. (2020). SciPy 1.0: Fundamental algorithms for scientific computing in Python. Nature Methods, 17(3), 261–272. 10.1038/s41592-019-0686-2

Weisz, N., Müller, N., Jatzev, S., & Bertrand, O. (2014). Oscillatory Alpha Modulations in Right Auditory Regions Reflect the Validity of Acoustic Cues in an Auditory Spatial Attention Task. Cerebral Cortex, 24(10), 2579–2590. 10.1093/cercor/bht113

Wen, D., Zhou, Y., & Li, X. (2015). A Critical Review: Coupling and Synchronization Analysis Methods of EEG Signal with Mild Cognitive Impairment. Frontiers in Aging Neuroscience, 7. 10.3389/fnagi.2015.00054

Wöstmann, M., Waschke, L., & Obleser, J. (2019). Prestimulus neural alpha power predicts confidence in discriminating identical auditory stimuli. European Journal of Neuroscience, 49(1), 94–105. 10.1111/ejn.14226

